# Plasma neurodegeneration biomarkers and clinical progression in early sporadic Parkinson’s disease

**DOI:** 10.64898/2026.11.18.26368420

**Authors:** Thomas F. Tropea, David G. Coughlin, Sarah M. Brooker, Krittika Chatterjee, Noah S. Johnson, Micah Marshall, Anuprita Nair, Javier E. Flores, David-Erick Lafontant, Christopher S. Coffey, Kristopher M. Kirmess, Tim West, Philip B. Verghese, Kristen A. Russ, Amanda Morris, Ming Lu, Leonardo Iaccarino, Patricia Aldea-Stevenson, Matthew Flitter, Rose Beck, Samantha Hutten, Lana M. Chahine, Daniel Weintraub, Tatiana Foroud, Tanya Simuni, Kenneth Marek, Kalpana Merchant, Parkinson Precision Medicine Initiative

## Abstract

**Objectives:** Plasma neurodegeneration biomarkers offer an opportunity to characterize Parkinson’s disease (PD) biology. The objective of this study was to investigate such biomarkers in deeply-phenotyped participants in Parkinson’s Precision Medicine Initiative (PPMI).

**Methods:** Baseline plasma samples from early (<2 years since diagnosis) sporadic PD participants with a positive CSF alpha-synuclein seed amplification assay (aSyn-SAA) and healthy controls (HC) with a negative aSyn-SAA test were utilized. Plasma P-tau217 (Roche Diagnostics), neurofilament light chain (NfL, Quanterix), glial fibrillary acidic protein (GFAP, Quanterix), and (PD only) amyloid-beta (Aβ) levels (40 and 42, C2N Diagnostics) were measured. Clinical features at baseline and over up to 11 years were compared across biomarker tertiles in models controlling for age, sex, and baseline outcome variable.

**Results:** The cohort was 97 HC and 523 PD and participants. Median [IQR] biomarker values (pg/mL) for HC and PD were P-tau217: 0.14 [0.06 -0.75], 0.15 [0.06 -0.90]; NfL: 10.6 [3.13 - 49.85], 12.26 [3.77-62.67]; GFAP: 66.56 [19.75 -403.48], 87.19 [14.55 -531.82]; and in PD Aβ40: 391.32 [209.95 -538.38]; Aβ42: 42.74[27.75 -68.55]. No biomarkers differed between HC and PD. In PD, biomarkers were associated with age. In PD only, higher baseline levels of P-tau217 and NfL were associated with greater hazards of a worse cognitive and balance outcome. P-tau217, NfL, and GFAP levels were associated with change in cognitive score over time.

**Interpretation:** Plasma P-tau217, NfL and GFAP levels in early, sporadic PD with confirmed synucleinopathy predict future cognitive and functional impairment, and these findings can inform clinical research design and clinical care.

## INTRODUCTION

Neuronal synuclein disease, which includes Parkinson’s disease (PD) and dementia with Lewy bodies (DLB), is defined by a positive alpha-synuclein seed amplification assay (aSyn-SAA). These overlapping neurodegenerative disorders are defined by presence of predominantly intraneuronal alpha-synuclein-containing Lewy body (LB) pathology and a spectrum of no clinical manifestations to motor and non-motor manifestations with increasing degrees of functional impairment.^1–3^

Significant heterogeneity exists in the clinical manifestation and progression of neuronal-predominant α-synucleinopathy, which may relate, in part, to the extent and anatomical distribution of LB pathology as well as the co-occurrence of other neuropathologies, including deposition of tau, amyloid and TDP-43 pathology, and occurrence of neuroinflammation. Co-occurring neurodegenerative pathologies or processes in α-synucleinopathies are clinically relevant and associate with faster progression and greater symptom burden. ^4–7^ Individuals with co-occurring tau and amyloid-beta (Aβ) in plaques and tangles characteristic of Alzheimer’s disease (AD) have a greater risk of cognitive impairment, earlier death, and greater symptom severity than those without AD co-pathology. ^8–14^ Based on an autopsy series, 30-50% of cases of confirmed PD cases have AD co-pathology at death.^9^

However, at earlier stages of the disease, closer to the time of diagnosis, the degree and impact of co-occurring neurodegenerative pathology is less well understood, in part because of the challenge posed by in vivo detection. Recent advances in the development of plasma biomarker assays offer an opportunity to characterize neurodegenerative processes and the detection of co-occurring brain pathology at early disease time points. ^15–18^

Blood-based markers of tau phosphorylated at threonine 217 (P-tau217) and amyloid-beta (Aβ) isoforms have comparable performance to cerebrospinal fluid (CSF) biomarkers and positron emission tomography (PET) imaging to detect brain Aβ pathology in cohorts of established PD. ^19–25^ Plasma glial fibrillary acidic protein (GFAP) is a non-specific marker of neuroinflammation that associates with AD co-pathology, ^18,26^ while neurofilament light chain (NfL) is a non-specific marker of axonal function/integrity. Both associate with disease progression in clinical cohorts of PD.^27,28^ Here we refer to these five markers collectively as biomarkers of neurodegeneration. Assessment of these biomarkers closer to diagnosis, and in individuals with confirmed CSF aSyn, could identify developing pathophysiology and its relationship with clinical progression to inform future therapeutic development ^29–31^.

The objectives of this study were to evaluate associations between neurodegeneration biomarker levels measured shortly after the clinical diagnosis in neuronal synuclein disease (NSD)-positive PD and symptom progression over time. We measured neurodegeneration biomarkers in baseline plasma from NSD+ sporadic PD cases and NSD-neurologically-healthy control (HC) participants aSyn enrolled in the Parkinson’s Precision Medicine Initiative (PPMI)^32^.

## METHODS

### Study Design and Participants

This case-control study follows the Strengthening of Reporting of Observational Studies in Epidemiology (STROBE) guidelines. Participants were enrolled in PPMI between 2010 and 2025 at 55 sites worldwide. PPMI is an international, longitudinal, observational study that collects clinical, biological, and imaging data from people with or at risk of PD and is registered at ClinicalTrials.gov (NCT01141023), with IRB approval at each site and written informed consent from participants. Full study details and enrollment criteria can be found at ppmi-info.org.^33^ Plasma and cerebrospinal fluid (CSF) samples were collected, processed, and stored at -80°C, as previously described. Methods can be found in the PPMI biologics manual at https://ppmi.info.org.

Participants were from one of two of the following cohorts:

- **Sporadic PD:** Inclusion required a PD clinical diagnosis within 2 years of enrollment, no prior PD medication use, Hoehn & Yahr stage 1–2, abnormal SPECT dopamine transporter scan (DaTscan), and at least two cardinal motor signs (with either resting tremor or bradykinesia required). Participants lacked PD relevant known pathogenic genetic variants (e.g., *LRRK2, GBA1, SNCA, PRKN,* etc.) All PD participants included for these analyses were CSF aSyn-SAA positive.
- **Healthy controls (HC)**: HC participants had no parkinsonism, dementia, or major neurological/psychiatric disease and a normal DaTscan at enrollment. All HC participants in this study were CSF aSyn-SAA negative.

Participants from the above cohorts were included in the baseline analysis if they had an available biomarker measurement and in the longitudinal analysis if they had at least one post-baseline visit.

### Clinical Outcomes

PPMI clinical outcomes include investigator-completed and participant-reported measures of motor, non-motor and cognitive function collected longitudinally; full details can be found at https://ppmi-online.org. Outcomes included: demographics (age, sex, race, ethnicity, education level, disease duration), Movement Disorder Society-Unified Parkinson’s Disease Rating Scale (MDS-UPDRS) Parts I-IV, Schwab & England activities of daily living score (S&E), age/sex-adjusted University of Pennsylvania Smell Identification Test (UPSIT) score, Hoehn & Yahr Score (HY), Scales for Outcomes in Parkinson’s disease-Autonomic (SCOPA-AUT) score, 15-item Geriatric Depression Scale (GDS-15) score, Montreal Cognitive Assessment (MoCA) score, and REM sleep behavior disorder (RBD) screening questionnaire (RBDSQ) score. Neuronal synuclein disease integrated staging system (NSD-ISS) stage was assigned at each visit as previously described.^3^ Cognitive diagnosis was made by the site investigator (normal cognition, mild cognitive impairment (MCI) or dementia). All assessments included in this analysis were conducted at baseline and at 6-12 month intervals.

We also analyzed time to reach domain-based disease milestones as previously described.^34^ Briefly, these include 25 progression milestones, spanning six clinical domains: “walking and balance”; “motor complications”; “cognition”; “autonomic dysfunction”; “functional dependence”; and “activities of daily living”.

### Biomarker Measurements

All biomarker studies were completed by qualified research staff who were blinded to clinical diagnosis and cohort assignment. Plasma biomarker measures were prioritized from study-entry samples when available; otherwise, samples collected within 6 months of study entry were used. These analyses were restricted to plasma biomarkers and included the following biomarkers: P-tau-217 was measured on the Roche Elecsys P-tau217 immunoassay using the Cobas® e 801 high throughput platform (Roche Diagnostics). Aβ40 and Aβ42 were measured by an immunoprecipitation-mass spectrometry (IP/MS, C2N Diagnostics) as described in Kirmess et al.^35^ GFAP and NfL were measured by the National Centralized Repository for Alzheimer’s Disease and Related Disorders (NCRAD) Biomarker Assay Laboratory on the Quanterix platform.^36^

CSF aSyn-SAA were measured by Amprion as previously described^37^ in samples collected at study entry. DaTscan was conducted as previously described in all participants at study entry (baseline) and at year 1 in 1129 participants.

### Statistical Analysis

Baseline demographic and clinical features were compared between PD and HC cohorts using two-sample test, chi-square test, or Fisher’s exact test, as appropriate. To account for differences due to age, linear regression and logistic regression adjusting for age were used to model clinical features and biomarkers with group as an explanatory variable. Log, square root, or rank transformations were applied to models with non-normally distributed residuals. The specific transformations used were marked on the tables and detailed in the table footers.

To define tertiles of each biomarker, PD and HC were combined with biomarker measurements from other cohorts within PPMI to derive the tertiles applicable across PPMI participants. **Supplemental Table 1** is a summary of all PPMI cohort biomarker measurements from which tertiles were assigned.

To assess whether the longitudinal trajectory of the outcome measures differed for different biomarker levels, generalized linear mixed-effects models (LMM) with random intercept and slope and unstructured working correlation structure were employed. Specifically, biomarker at baseline (both as continuous measure and as tertiles), time in years, and their interaction were included in the models. Wald tests were conducted to assess the statistical significance of the interaction term between biomarker levels and time. All longitudinal models adjusted for baseline value of the outcome, age, sex, and were conducted under the assumption of missing at random (MAR), which was evaluated with the Index of Local Sensitivity to Non-Ignorability (ISNI).^38^

Models included age and sex as covariates. Education has minimal variability in this cohort and was not included in analyses as a covariate. *APOE* genotype was evaluated, though with minimal impact on overall outcomes (indicated in results) and is not included in final models. Given the exploratory nature of this study, we set the significance threshold at 0.05 (two-tailed). All analyses were conducted in SAS Institute Inc. (SAS Institute Inc version 9.4 Cary, NC).

## RESULTS

### Participant Cohorts

523 PD participants met criteria to be a part of this study (48% of total PD enrollees in PPMI n=1079 as of 8th December 2025). 97 HC participants met criteria for this study (29% of HC participants in PPMI study). Baseline demographic and clinical characteristic for PD and HC cohorts are summarized in **Table 1**.

**Table 1:** Baseline demographic and clinical characteristic.

|  | Healthy Control<br>**NSD-**<br>(N = 97) | Sporadic PD<br>**NSD+**<br>(N = 523) | **Pairwise p-values**<br>**HC vs sPD** |
|---|---|---|---|
| **Sex**, n (%) |  |  |  |
| Male | 57 (59%) | 323 (62%) | 0.5779 |
| Female | 40 (41%) | 200 (38%) |  |
| **Age (years)**, mean (SD) | 58.9 (12.4) | 62.9 (9.3) | **0.0041** |
| Median (min, max) | 58.4 (30.6, 81.3) | 63.7 (33.7, 84.9) |  |
| **Disease Duration (years)**, mean (SD) | NA | 0.7 (0.6) | NA |
| Median (min, max) | NA | 0.5 (0.0, 3.2) |  |
| **Race**, n (%) |  |  |  |
| White | 88 (91%) | 485 (93%) | 0.3708 |
| Black/African American | 6 (6%) | 8 (2%) |  |
| Asian | 1 (1%) | 10 (2%) |  |
| Other | 2 (2%) | 17 (3%) |  |
| Missing | 0 | 3 |  |
| **Revised UPSIT percentile**, mean (SD) | 59.9 (28.5) | 10.9 (13.3) | **<.0001** |
| Median (min, max) | 62.5 (2.0, 100.0) | 6.0 (1.0, 91.5) |  |
| **Revised UPSIT percentile ≤ 15**, n (%) | 10 (10%) | 403 (79%) | **<.0001** |
| **NSD+**, n (%) |  |  | NA |
| 2a | NA | 8 (2%) |  |
| 2b | NA | 74 (14%) |  |
| 3 | NA | 409 (78%) |  |
| 4 | NA | 32 (6%) |  |
| 5 | NA | 0 |  |
| Missing | NA | 0 |  |
| **MDS-UPDRS item 1.1**, n (%) |  |  | **0.0002** <sup>a</sup> |
| Normal | 91 (95%) | 387 (74%) |  |
| Slight | 4 (4%) | 125 (24%) |  |
| Mild | 1 (1%) | 11 (2%) |  |
| Moderate | 0 | 0 |  |
| Missing | 1 | 0 |  |
| **MDS-UPDRS part I**, mean (SD) | 2.7 (2.8) | 5.7 (3.7) | **<.0001** <sup>c</sup> |
| Median (min, max) | 2 (0, 12) | 5 (0, 23) |  |
| Missing | 1 | 0 |  |
| **MDS-UPDRS part II**, mean (SD) | 0.6 (1.1) | 6.0 (3.7) | **<.0001** <sup>c</sup> |
| Median (min, max) | 0 (0, 6) | 5 (0, 22) |  |
| Missing | 1 | 0 |  |
| **MDS-UPDRS part III**, mean (SD) | 1.0 (2.1) | 22.3 (9.3) | **<.0001** <sup>c</sup> |
| Median (min, max) | 0 (0, 13) | 21 (4, 52) |  |
| Missing | 1 | 0 |  |
| **MDS-UPDRS total score**, mean (SD) | 4.2 (4.3) | 34.0 (12.7) | **<.0001** <sup>b</sup> |
| Median (min, max) | 3 (0, 17) | 33 (7, 70) |  |
| Missing | 1 | 0 |  |
| **MoCA total score**, mean (SD) | 28.4 (1.2) | 27.1 (2.3) | **<.0001** <sup>c</sup> |
| Median (min, max) | 28 (27, 30) | 28 (17, 30) |  |
| Missing | 0 | 0 |  |
| **SCOPA-AUT total score**, mean (SD) | 5.7 (3.7) | 9.7 (5.8) | **<.0001** <sup>c</sup> |
| Median (min, max) | 5 (0, 18) | 9 (0, 39) |  |
| Missing | 0 | 6 |  |
| **Hoehn and Yahr**, n (%) |  |  | 0.9147<sup>a</sup> |
| Stage 0 | 95 (99%) | 0 |  |
| Stage 1 | 1 (1%) | 198 (38%) |  |
| Stage 2 | 0 | 325 (62%) |  |
| Stage 3 | 0 | 0 |  |
| Missing | 1 | 0 |  |
| **% Age/sex-expected low putamen**, mean (SD) | 1.00 (0.15) | 0.41 (0.14) | **<.0001** |
| Median (min, max) | 1.00 (0.65, 1.35) | 0.40 (0.02, 0.99) |  |
| Missing | 2 | 0 |  |
| **Cognitive Categorization**, n (%) |  |  | 0.9751<sup>a</sup> |
| Normal cognition | 15 (100%) | 245 (93%) |  |
| Mild Cognitive Impairment | 0 | 19 (7%) |  |
| Missing | 82 | 259 |  |

<sup>a</sup> P-values calculated using PROC LOGISTIC adjusted for age and sex. <sup>b</sup> P-values calculated using a square-root-transformed model. <sup>c</sup> P-values calculated using a rank-based nonparametric model. No superscript indicates unadjusted p-values. Race p-value is a comparison of non-white vs. white. For MDS-UPDRS item 1.1, all groups except 'Normal' were collapsed and the resulting p-value is a comparison of normal vs. any pain between two groups For NSD Stage, Stages <2B are collapsed and compared against Stages 3<. For Hoehn and Yahr, Stage 0 is compared against Stages >0.

Participants were followed annually for up to 11 years, with a median (min-max) follow up of 5 years (2-11) in PD, and 10 years (5-11) in HC participants. The number of participants followed at each yearly interval for each group is shown in **Supplemental Figure 1**.

### Summary of Plasma Biomarker Samples

620 samples (all PD and HC participants) were included in these analyses, contributing 1,147 biomarker samples across the assays. Across biomarkers, 1,040 samples were from the baseline/study-entry visit (846 PD and 194 HC), and 107 samples were from within 6 months of study entry (107 PD and 0 HC). Baseline and 3M/6M sample counts were P-tau217: 1,292 BL and 143M/6M; NfL and GFAP: 1,578 BL and 163M/6M; Aβ40 and Aβ42: 70 BL and 163 3M/6M. P-tau217 was measured in 428 samples (331 PD and 97 HC), GFAP and NfL in 619 samples (522 PD and 97 HC), and Aβ42 and Aβ40 in 100 samples (100 PD and 0 HC). Results were missing due to inconclusive assay measurements in 8 samples (all 8 were in P-tau217 falling under lower limit of detection (LLOD, 0.05). No differences in biomarker levels or the frequency of cases within each tertile were detected between PD and HC groups with the exception of GFAP, which differed on the frequency of cases within each tertile (χ^2^ (1) =4.68, p= 0.0305) with 23% of HC and 36% of PD in the highest tertile. **Table 2** and **Supplemental Figure 2** are summaries of biomarker measures within PD and HC cohorts.

**Table 2:** Baseline plasma biomarkers.

|  | **Healthy Control**<br>**NSD-**<br>(N = 97) | **Sporadic PD**<br>**NSD+**<br>(N = 523) | **p-values**<br>**HC vs sPD** |
|---|---|---|---|
| **Ptau217**, mean (SD) | 0.17 (0.10) | 0.18 (0.10) | 0.5687<sup>c</sup> |
| Median (min, max) | 0.14 (0.06, 0.75) | 0.15 (0.06, 0.90) |  |
| Missing | 1 | 200 |  |
| **Ptau217 tertiles**, n (%) |  |  | 0.4192<sup>a</sup> |
| < 0.131 | 40 (41%) | 109 (33%) |  |
| 0.131 - 0.174 | 30 (31%) | 112 (34%) |  |
| > 0.174 | 27 (28%) | 110 (33%) |  |
| Missing | 0 | 192 |  |
| **NFL**, mean (SD) | 12.03 (7.15) | 13.52 (6.24) | 0.2445<sup>c</sup> |
| Median (min, max) | 10.60 (3.13, 49.85) | 12.26 (3.77, 62.67) |  |
| Missing | 0 | 1 |  |
| **NFL tertiles**, n (%) |  |  | 0.0695<sup>a</sup> |
| < 9.877 | 44 (45%) | 172 (33%) |  |
| 9.877 - 14.538 | 33 (34%) | 163 (31%) |  |
| > 14.538 | 20 (21%) | 187 (36%) |  |
| Missing | 0 | 1 |  |
| **GFAP**, mean (SD) | 84.76 (58.45) | 98.70 (55.51) | 0.1372<sup>b</sup> |
| Median (min, max) | 66.56 (19.75, 403.48) | 87.19 (14.55, 531.82) |  |
| Missing | 0 | 1 |  |
| **GFAP tertiles**, n (%) |  |  | **0.0305<sup>a</sup>** |
| < 64.478 | 45 (46%) | 144 (28%) |  |
| 64.478 - 105.341 | 30 (31%) | 191 (37%) |  |
| > 105.341 | 22 (23%) | 187 (36%) |  |
| Missing | 0 | 1 |  |
| **Aß40**, mean (SD) | NA | 387.67 (59.12) | NA |
| Median (min, max) | NA | 391.32 (209.95, 538.38) |  |
| Missing | 97 | 423 |  |
| **Aß40 tertiles**, n (%) |  |  | NA |
| < 374.886 | 0 | 37 (37%) |  |
| 374.886 - 422.785 | 0 | 40 (40%) |  |
| > 422.785 | 0 | 23 (23%) |  |
| Missing | 97 | 423 |  |
| **Aß42**, mean (SD) | NA | 43.31 (7.54) | NA |
| Median (min, max) | NA | 42.74 (27.75, 68.55) |  |
| Missing | 97 | 423 |  |
| **Aß42 tertiles**, n (%) |  |  | NA |
| < 40.219 | 0 | 32 (32%) |  |
| 40.219 - 45.870 | 0 | 37 (37%) |  |
| > 45.870 | 0 | 31 (31%) |  |
| Missing | 97 | 423 |  |
| **Aß42/ Aß40**, mean (SD) | NA | 0.11 (0.02) | NA |
| Median (min, max) | NA | 0.11 (0.09, 0.20) |  |
| Missing | 97 | 423 |  |
| **Aß42/ Aß40 tertiles**, n (%) |  |  | NA |
| < 0.104 | 0 | 26 (26%) |  |
| 0.104 - 0.111 | 0 | 29 (29%) |  |
| > 0.111 | 0 | 45 (45%) |  |
| Missing | 97 | 423 |  |

<sup>a</sup> P-values calculated using PROC LOGISTIC adjusted for age and sex. <sup>b</sup> P-values calculated using a log-transformed model. <sup>c</sup> P-values calculated using a rank-based nonparametric model. P-values for biomarker cutoffs were calculated using a cumulative logit link function.

### Associations between biomarker levels and baseline clinical assessments

For both PD and HC subgroups, age was directly associated with higher tertiles of P-tau217 (PD:[χ^2^(2)=29.31], p<0.0001, HC: χ^2^(2)=7.74, p=0.0209), NfL (PD:[χ^2^(2)=203.00], p<0.0001, HC: [χ^2^(2)=51.48], p<0.0001), GFAP (PD: [χ^2^(2)=149.75], p<0.0001, HC: [χ^2^(2)=29.63], p<0.0001), and Aβ40 (PD: [χ^2^(2)=19.87], p<0.0001), and indirectly with Aβ42 (PD: [χ^2^(2)=8.25], p=0.0162), and Aβ42/40 (PD: [χ^2^(2)=7.58], p =0.0226). For the PD group, P-tau217 ([χ^2^(2)=6.46], p=0.0395) and GFAP ([χ^2^(2)=8.55], p=0.0139) differed by sex, but both PD and HC subgroups indicated a similar trend of a larger proportion of males in the middle tertile versus higher and lower tertiles.

Among PD cases, higher P-tau217 values were associated with worse scores on rank-transformed MDS-UPDRS Part II (β=117.026, 95% CI [9.297,224.755], p=0.0333) and rank-transformed SCOPA-AUT (β=109.310, 95% CI [3.733,214.887], p=0.0425). Lower DaTscan mean lowest putamen ratio percentile was associated with elevations in both NfL (β=0.0031, 95% CI [0.0012,0.0050], p=0.0015) and GFAP (β=0.0003, 95% CI [<0.0001,0.0005], p=0.0234).

Additionally, associations between lower levels of Aβ40 and log-transformed MDS-UPDRS Part I (β=-0.003, 95% CI [-0.006, -0.0004], p=0.0260) and log-transformed MDS-UPDRS Part II scores (β=-0.003, 95% CI [-0.006, -0.0002], p=0.0357) were identified. There were no differences in clinical outcome measures across biomarker tertiles in the HC group. Results for each biomarker for all clinical outcome measures at baseline are shown in **Supplemental Tables 2a-i**.

### Associations between biomarker level and longitudinal cognitive outcomes

In PD, faster decline in MoCA score over time was associated with greater levels of P-tau217 [Est. (95% CL): -1.787 (-2.807,-0.767), p=0.0006], NfL (-0.015 (-0.028,-0.002), p=0.0235] and GFAP (: -0.002(-0.004,-0.001), p=0.0027), but not Ab42 (p=0.9953), Ab40 (p=0.6848), or Ab42/40 (p=0.5856). Higher P-tau217 [HR(95% CL): 23.887 (5.458,104.544), p<0.001] and GFAP [HR(95% CL): 1.004 (1.001,1.007), p=0.016], but not NfL (p=0.268), Ab42 (p=0.468), Ab40 (p=0.704), or Ab42/40 (p=0.318), were associated with a greater hazard of achieving an impaired cognition milestone (**Figure 1).** In models covarying for APOE genotype, the association between GFAP and cognitive diagnosis change was no longer significant (p=0.061), while all other results were unaffected.

**Figure 1.**
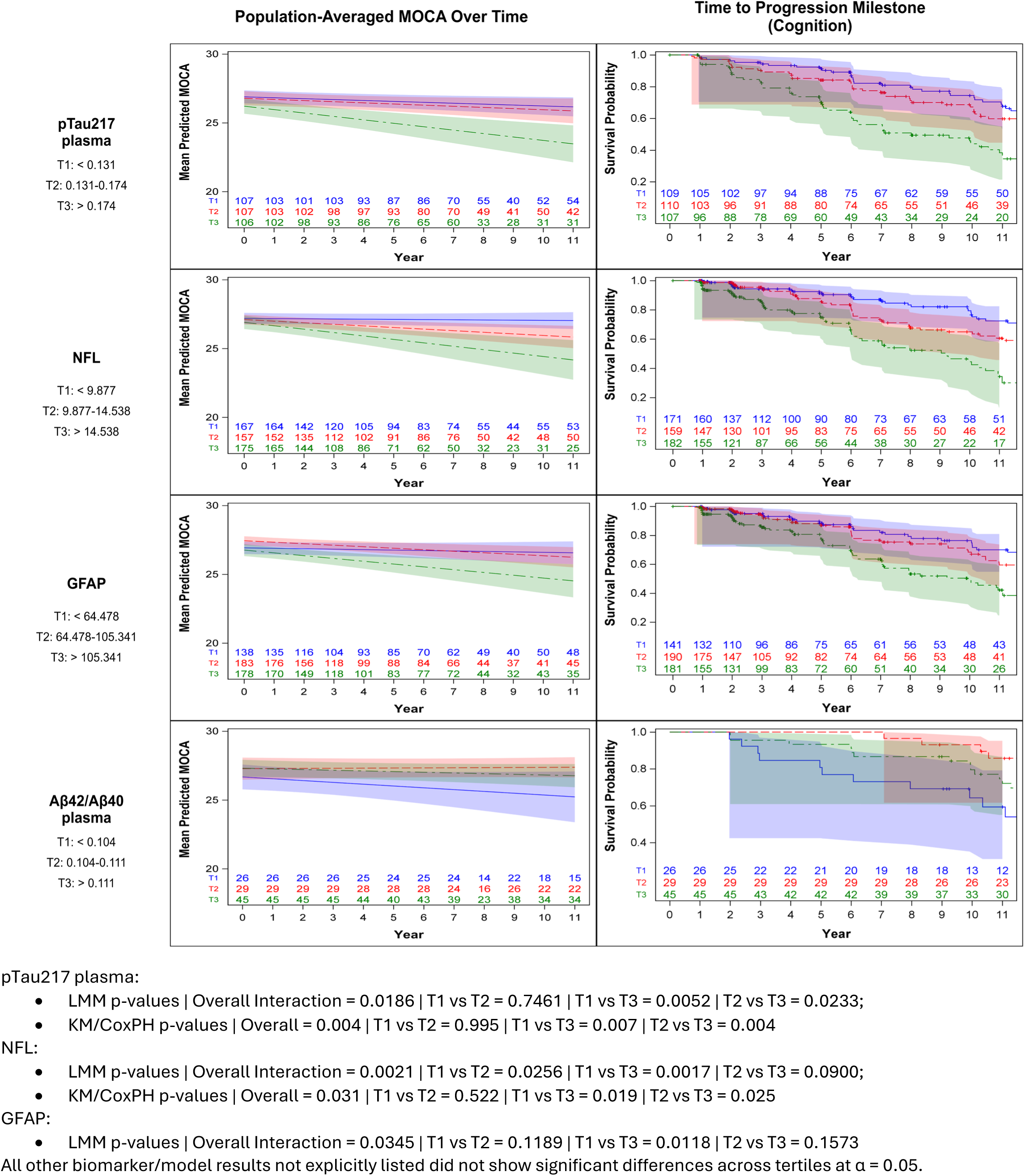
Plasma P-tau217 and NfL concentration in Parkinson’s disease participants predicts cognitive progression. Top row left, row 2 left, row 3 left: P-tau217, NfL, and GFAP concentration measured at study entry predicts yearly rate of change in the Montreal Cognitive Assessment (MoCA) in participants followed for up to 11 years. Top right and row 2 right: Risk of meeting a cognitive progression milestone (defined by Brumm et al) differed by P-tau217 or NfL concentration and individuals in the highest tertiles of either P-tau217 (>0.174 pg/mL) or NfL (>14.538 pg/mL) showed greater risk compared to the lower tertiles.

In the HC cohort, no associations between baseline biomarker and MoCA progression was detected for P-tau217, GFAP, or NfL, but higher P-tau217 did predict a greater hazard for achieving a MoCA score MoCA<26 (HR(95%CL): 38.01 (3.15, 458.17), p=0.004). In HC, only two progression milestone events occurred, so time-to-event analyses were not conducted. See **Supplemental Table 3a-b (biomarker as continuous measure)** and **Supplemental Table 3c-d (biomarker by tertile)** for summaries of mixed-effects models evaluating associations between biomarker and clinical outcome measures. See **Supplemental Table 4a-c** (biomarker as continuous measure) and **Supplemental Table 4d-f** (biomarker by tertile) for all time-to-event analyses for clinical measures and progression milestones for all clinical outcome measures in PD and HC cohorts.

### Associations between biomarker level and longitudinal non-motor outcomes

In PD, higher levels of P-tau217 [Est. (95% CL): -1.808 (0.320,3.296), p=0.0173] and NfL [Est. (95% CL): 0.022 (0.003,0.042), p=0.0233], but not GFAP (p= 0.1095), Ab42(p=0.2878), Ab40 (p=0.9872) or Ab42/40 (p=0.1056), predicted a faster rate of increase in MDS-UPDRS Part I score. Cox proportional hazards models were used to assess the relationships between biomarker and risk of autonomic composite endpoint and accelerated failure time (AFT) model was used for functional dependence endpoint with covariates of age and sex. Higher P-tau217, but not other biomarkers, associated with a greater hazard of meeting a functional dependence milestone [HR(95% CL): 14.126 (3.051,65.397), p<0.001]. In models covarying for APOE genotype, the association between GFAP and autonomic dysfunction became significant (p=0.046), while all other results were unaffected. **Figure 2** shows associations between baseline biomarkers and non-motor outcomes in the PD cohort across tertiles of biomarker level.

**Figure 2.**
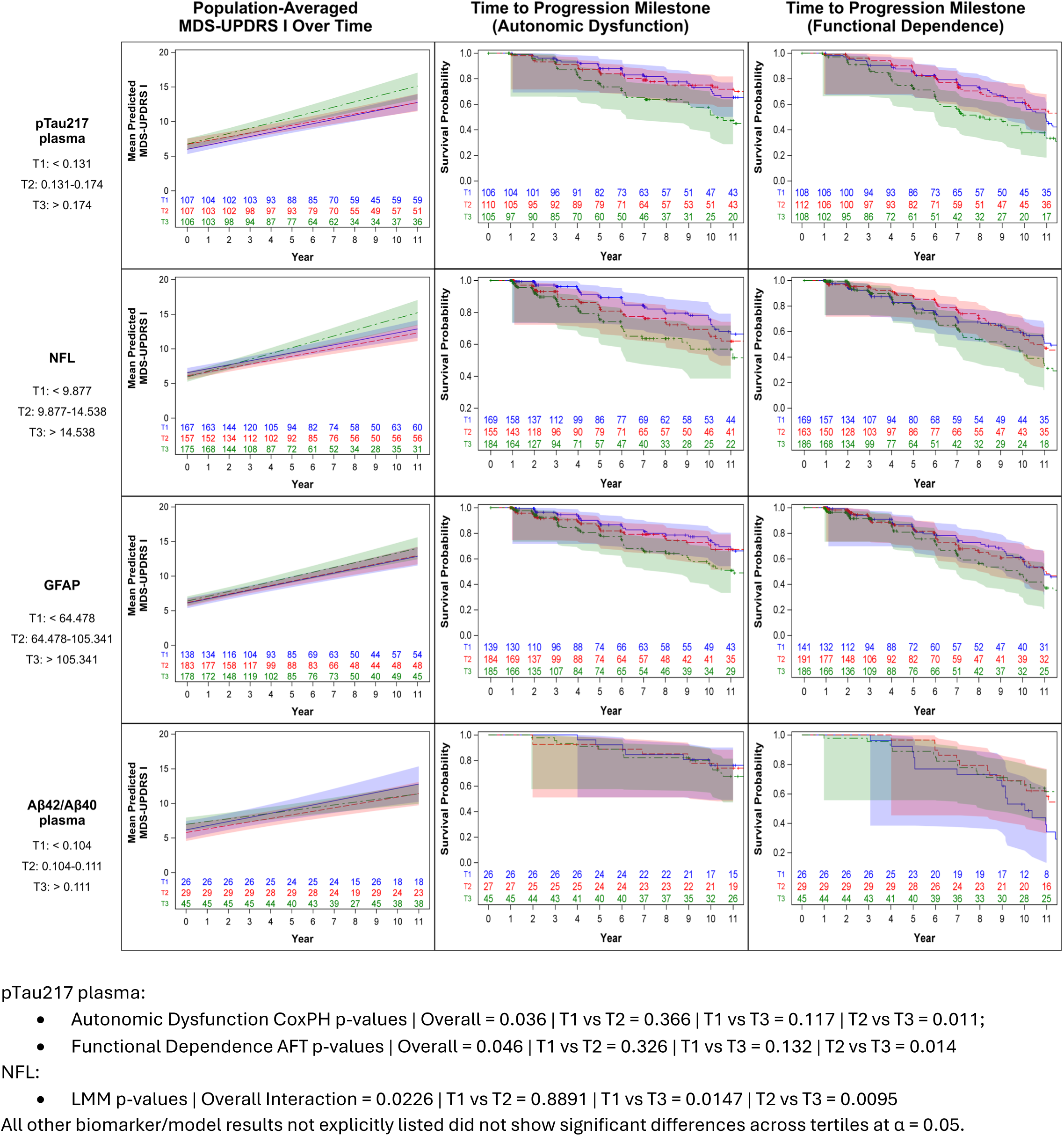
Associations between baseline biomarkers and non-motor outcomes in the PD cohort across tertiles of biomarker level. Top-middle and Top-right: Risk of meeting an autonomic or functional dependence outcome (defined by Brumm et al) differed by P-tau217 concentration and individuals in the highest tertiles of P-tau217 (>0.174 pg/mL) showed greater risk compared to the lower tertiles. Left row 2: NfL concentration measured at study entry predicts yearly rate of change in the Movement Disorders Society Unified Parkinson’s Disease Rating Scale Part 1 Total Score in participants followed for up to 11 years.

In the HC cohort, no associations between baseline biomarker and the above outcome measures were observed, and no events occurred and thus time-to-event analyses were not conducted. See **Supplemental Table 3a-d** and **Supplemental Table 4a-f** for a summary of mixed-effects model outcomes and for all time-to-event analyses for all clinical outcome measures in PD and HC cohorts.

### Associations between biomarker level and longitudinal motor outcomes

In PD, linear mixed-effects models, adjusting for age, sex, and baseline score, showed that elevations in P-tau217 [Est. (95% CL): 2.6437(0.5286,4.7588), p=0.0143] and NfL [Est. (95% CL): 0.0281(0.0046,0.0517), p=0.0191], but not GFAP (p= 0.0565), Ab42(p=0.8179), Ab40 (p=0.3192), or Ab42/40 (0.0575), predicted a faster rate of increase in MDS-UPDRS Part II scores over time. Lower Ab42/40 was associated with faster rate of increase in MDS-UPDRS Part III OFF; [Est. (95% CL): -15.8612(-29.7053,-2.0171), p=0.0248], but no other associations were detected in PD or HC cohorts. Cox proportional hazards models assessed relationships between biomarker and risk of motor complications, and AFT model was used for walking and balance endpoints, with covariates of age and sex but no associations were detected in the PD cohort. Very few or no events occurred in the HC cohort, so an analysis wasn’t conducted. **Figure 3** shows associations between baseline biomarkers and motor outcomes in the PD cohort across tertiles of biomarker level. See **Supplemental Table 3a-d** and **Supplemental Table 4a-f** for a summary of mixed-effects model outcomes for all clinical outcome measures.

**Figure 3.**
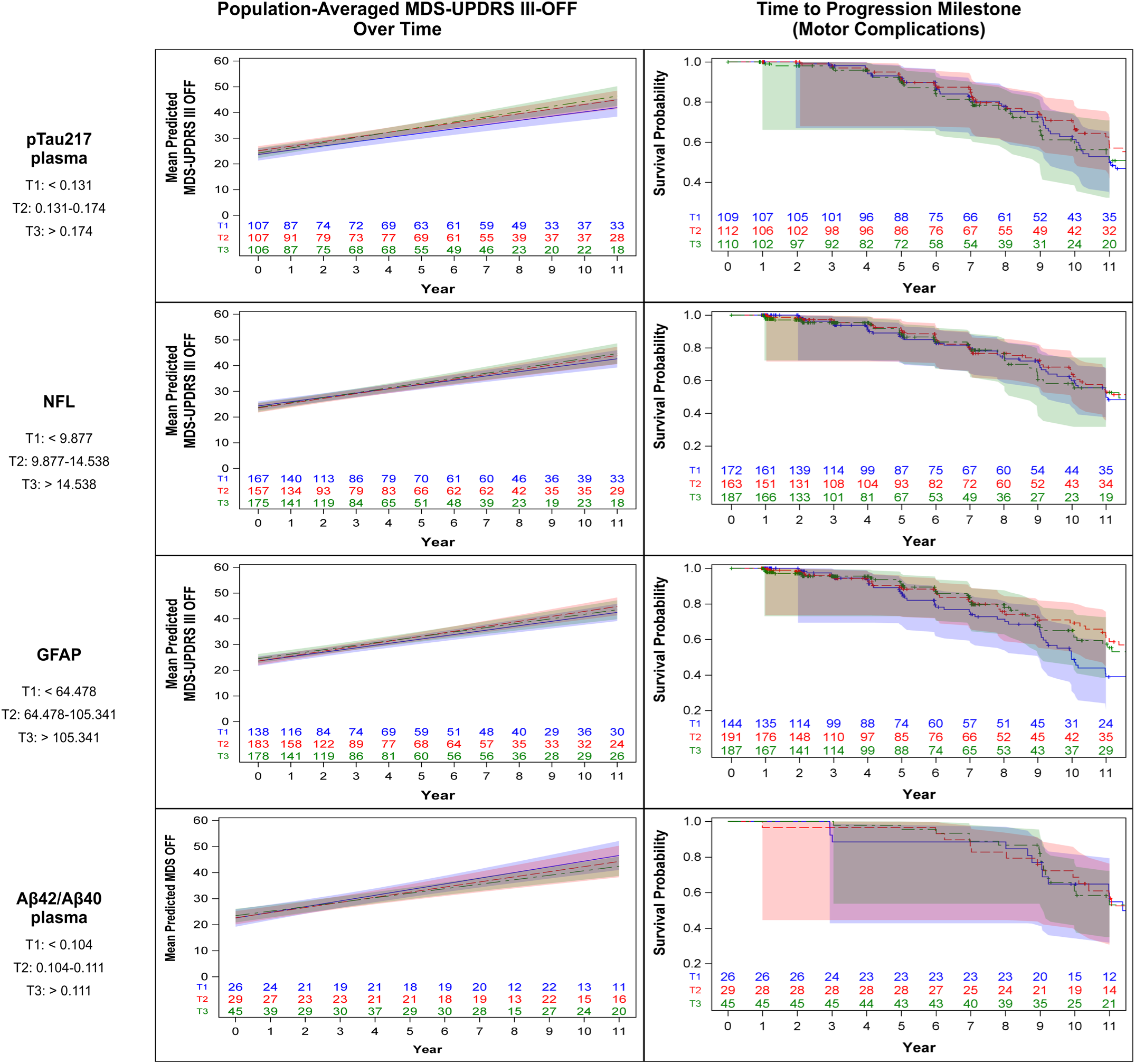
Associations between baseline biomarkers and motor outcomes in the PD cohort across tertiles of biomarker level. P-tau217, NfL, or GFAP concentration or Amyloid-Beta 42/40 ratio did not predict change in the Movement Disorders Society Unified Parkinson’s Disease Rating Scale Part 3 Total Score or risk of reaching a motor progression milestone (defined by Brumm et al).

### Associations between biomarker level and subsequent DaTscan changes

No associations between any of the biomarkers measured and changes in DaTscan mean lowest putamen ratio over time over 4 years of follow-up were observed. See **Supplemental Table 3a-d** and **Supplemental Table 4a-f** for a summary of mixed-effects model outcomes for all clinical and imaging outcome measures.

## DISCUSSION

We examined associations between baseline plasma neurodegenerative disease biomarkers and cross-sectional and longitudinal clinical variables in a deeply-phenotyped cohort of aSyn-SAA positive individuals with recently diagnosed sporadic PD and a comparator group of aSyn-SAA negative HC. This work shows that such biomarkers, particularly p-tau217, in recently diagnosed PD associates with longitudinal disease progression for cognitive and other functional outcomes. Additionally, biomarker measures were higher in older individuals, although there were no differences between PD and HC cohorts. These findings demonstrate that plasma neurodegeneration biomarkers in early disease states of aSyn-SAA positive PD predict cognitive progression and functional decline but not DaTscan changes.

Plasma biomarkers have become central to the diagnosis of dementia due to AD. In this context, increased levels of plasma biomarkers, including P-tau and Aβ isoforms, are detectable years prior to cognitive decline, representing early states of the neurodegenerative cascade ultimately leading to dementia and functional decline secondary to amyloid and tau pathology. In PD cohorts, antemortem plasma measures of P-tau isoforms associate with AD co-pathology in autopsy cases.^16,18,39,40^ Additionally, P-tau isoforms predict AD co-pathology in earlier disease when compared with amyloid PET and CSF-based biomarkers as the gold standard.^25^

Yet, measures of P-tau biomarkers in PD may not reflect only established amyloid or tau neuropathology. In studies of PD, the magnitude of change in Aβ and tau biomarkers reflect the burden of underlying AD co-pathology and tends to be lower in magnitude than in cohorts of patients with dementia due to AD.^18,39,41,42^ The levels of P-tau217 seen in our study are lower relative to cutoffs established in AD despite their associations with clinical progression. This may indicate that lower levels of underlying amyloid pathology or other pathophysiologic mechanism, reflected as lower P-tau217 levels, are clinically relevant and predict disease progression. Indeed, despite their consistent relationship with worse outcomes, pathology studies in PD show that the magnitude and extent of AD co-pathology in PD is commonly less severe in density and distribution than in AD suggesting that a lower burden of these pathologies may be harmful when found in combination compared to when found in isolation.

Alternatively, circulating plasma P-tau217 in PD may be indicative of an underlying pathophysiologic process of copathology development. *In vitro* studies have demonstrated interactions between aSyn and tau leading to protein misfolding and aggregation. Elevated levels of circulating P-tau isoforms may enhance aSyn propagation leading to greater aSyn misfolding and deposition alongside AD co-pathology. Additional work is needed to better characterize the early development of tau and amyloid pathology.

Beyond P-tau isoforms, associations between plasma GFAP, which is a non-specific marker of neuroinflammation and gliosis, and AD co-pathology have been reported in synucleinopathies. Indeed, in one report, antemortem plasma GFAP better predicted AD co-pathology in PD and DLB cases compared with plasma P-tau181. In clinical cohorts of PD, higher level of GFAP was associated with worse cognitive function. Small studies have demonstrated marginal associations of GFAP with longitudinal cognitive decline.^26,43,44^ Elevations in GFAP, reflective of astrogliosis and neuroinflammation leading to and/or resulting from a greater occurrence of AD co-pathology, sheds light on the pathophysiologic process in early-stage PD.

Furthermore, elevations in NfL that may reflect severity of neurodegeneration,^45^ have been reported in association in PD, although these associations are modest and not universally reported. A number of studies have reported associations between plasma NfL and motor and cognitive progression.^28,46–48^ Furthermore, greater increases in serially-measured serum NfL have been shown to associate with greater risk of cognitive impairment.^27^ While our work does not show an association between NfL and cognitive progression, there is an association with non-motor functional decline. Taken together, the severity of neuroinflammation and neurodegeneration at early disease timepoints may indicate a more aggressive form of PD with greater risk of functional decline over time.

There are important caveats to consider when interpreting these data. For one, there is incomplete overlap between individuals with different plasma biomarkers due to selection of samples for different analytes. The selection criteria of each group, however, was not different and was based on availability of samples and scope of each analyte project. Importantly, the HC group was not included in the Ab study, which was due to the smaller scope of that study.

Second, while these cases are extensively clinically characterized, they lack tau or amyloid PET imaging characterization, which limits our interpretation of their comparable plasma biomarkers. Ongoing studies are obtaining tau and amyloid PET data alongside additional plasma samples for future biomarker-imaging association studies. Third, in the absence of defined cutoffs of these biomarkers relevant in PD specifically, tertiles across the entire study population were used to group individuals for analysis. The interpretation of those tertiles is not intended to define new cutoffs, but rather to identify trends in the clinical data across biomarker groups. Future studies to validate amyloid and tau biomarker cutoffs in PD to predict AD co-pathology are needed. Finally, while we demonstrate biomarkers in an extensively-phenotyped cohort with long-term follow up, an appropriate validation cohort does not exist, limiting the interpretation and application of these findings.

With these important considerations, we highlight some strengths of this work that may underlie the differences in our work compared with other reports. For one, all participants in this study are enrolled within 2 years of a clinical diagnosis of PD and followed longitudinally, and biomarkers were measured at these early time points. Furthermore, all participants have known aSyn-SAA status. All PD included were aSyn-SAA positive and genetic PD groups are excluded from this study, reducing pathological heterogeneity. Such depth in biological and clinical characterization is a major strength of this work.

## Conclusion

Elevated levels of plasma neurodegeneration biomarkers, particularly P-tau217, in sporadic PD with confirmed CSF alpha-synuclein positivity, predict longitudinal cognitive and broader functional decline, but not dopaminergic imaging outcomes. Taken together, these findings indicate that P-tau217 is a promising prognostic biomarker in recently diagnosed PD that may be informative to assess for enrichment or stratification of cohorts in clinical trials of disease-modifying therapies.

## Supporting information

PPMI Study Group Authors

## Author Contributors

TFT, DGC, SMB, TS, KM, and KMa contributed to the conception and design of the study. TFT, KC, NSJ, MM, AN, JEF, DEL, and CSC contributed to the analysis of the data. All authors contributed to drafting or reviewing a significant portion of the manuscript or figures.

## Data Availability

Data used in the preparation of this article were obtained on 2025-12-08 from the Parkinson’s Precision Medicine Initiative (PPMI) database (https://www.ppmi-info.org/access-data-specimens/download-data), RRID:SCR_006431. For up-to-date information on the study, visit http://www.ppmi-info.org.

This analysis was conducted by the PPMI Statistics Core and used actual dates of activity for participants, a restricted data element not available to public users of PPMI data.

Protocol information for The Parkinson’s Precision Medicine Initiative (PPMI) Clinical - Establishing a Deeply Phenotyped PD Cohort can be found on protocols.io or by following this link: https://dx.doi.org/10.17504/protocols.io.n92ldmw6ol5b/v2.

Statistical analysis codes used to perform the analyses in this article are shared on GitHub [https://github.com/UI-CTSDMC/Plasma_Biomarkers_Analysis] and were given a persistent identifier via Zenodo [10.5281/zenodo.23221473].

PPMI – a public-private partnership – is funded by the Michael J. Fox Foundation for Parkinson’s Research and funding partners, including AbbVie, Alamar Biosciences, Aligning Science Across Parkinson’s (ASAP), Arrowhead Pharma, Arvinas, AskBio, BIAL, BioArctic, Biohaven, BlueRock Therapeutics, Bristol Myers Squibb, Calico Labs, Capsida Biotherapeutics, Critical Path Institute, DaCapo Brainscience, Denali, Edmond J. Safra Foundation, Eli Lilly, Gain Therapeutics, GE Healthcare, Genentech, GSK, Insitro, Johnson & Johnson Innovative Medicine, Lundbeck, Merck, Neumora, Neuron23, Novartis, Olink, Regeneron, Roche, Sanofi, Tenvie, UCB, Vanqua Bio, Voyager Therapeutics, The Weston Family Foundation.

The Biomarker Assay Lab is supported by the National Centralized Repository for Alzheimer’s Disease and Related Dementias (NCRAD) through a cooperative agreement grant (U24 AG021886) awarded to NCRAD by the National Institute on Aging (NIA).

