## Supplement Materials for "Plasma neurodegeneration biomarkers and clinical progression in early sporadic Parkinson’s disease"

#### Supplemental Table 1: Summary of All PPMI Cohort Biomarker Measurements

| **Variable** | **Overall** (N=860) | **Sporadic PD** (N=551) | **Healthy Control** (N=100) | **LRRK2 PD** (N=142) | **GBA PD** (N=67) |
| --- | --- | --- | --- | --- | --- |
| **pTau217**, mean (SD) | 0.18 (0.10) | 0.18 (0.10) | 0.17 (0.10) | 0.17 (0.08) | 0.19 (0.13) |
| Median (min, max) | 0.15 (0.06, 0.90) | 0.15 (0.06, 0.90) | 0.14 (0.06, 0.75) | 0.15 (0.06, 0.55) | 0.16 (0.06, 0.82) |
| Missing | 298 | 221 | 1 | 60 | 16 |
| **pTau217 tertiles**, n (%) |  |  |  |  |  |
| < 0.131 | 203 (35%) | 111 (33%) | 40 (40%) | 32 (38%) | 20 (36%) |
| 0.131 - 0.174 | 190 (33%) | 115 (34%) | 31 (31%) | 26 (31%) | 18 (33%) |
| > 0.174 | 185 (32%) | 113 (33%) | 29 (29%) | 26 (31%) | 17 (31%) |
| Missing | 282 | 212 | 0 | 58 | 12 |
| **NFL**, mean (SD) | 13.49 (6.77) | 13.62 (6.26) | 12.20 (7.16) | 13.86 (7.30) | 13.58 (8.74) |
| Median (min, max) | 12.16 (3.13, 66.50) | 12.31 (3.77, 62.67) | 10.75 (3.13, 49.85) | 12.43 (4.79, 60.46) | 11.53 (4.63, 66.50) |
| Missing | 2 | 1 | 0 | 0 | 1 |
| **NFL tertiles**, n (%) |  |  |  |  |  |
| < 9.8771 | 285 (33%) | 176 (32%) | 44 (44%) | 38 (27%) | 27 (41%) |
| 9.8771 - 14.5384 | 288 (34%) | 175 (32%) | 34 (34%) | 58 (41%) | 21 (32%) |
| > 14.5384 | 285 (33%) | 199 (36%) | 22 (22%) | 46 (32%) | 18 (27%) |
| Missing | 2 | 1 | 0 | 0 | 1 |
| **GFAP**, mean (SD) | 95.09 (55.29) | 98.53 (55.31) | 86.00 (58.98) | 88.98 (47.17) | 93.35 (63.52) |
| Median (min, max) | 81.66 (14.55, 531.82) | 86.56 (14.55, 531.82) | 67.09 (19.75, 403.48) | 75.60 (23.47, 248.64) | 70.48 (16.59, 375.41) |
| Missing | 2 | 1 | 0 | 0 | 1 |
| **GFAP tertiles**, n (%) |  |  |  |  |  |
| < 64.4776 | 285 (33%) | 154 (28%) | 46 (46%) | 57 (40%) | 28 (42%) |
| 64.4776 - 105.3412 | 288 (34%) | 200 (36%) | 30 (30%) | 44 (31%) | 14 (21%) |
| > 105.3412 | 285 (33%) | 196 (36%) | 24 (24%) | 41 (29%) | 24 (36%) |
| Missing | 2 | 1 | 0 | 0 | 1 |
| **Aβ40**, mean (SD) | 408.11 (85.34) | 386.94 (58.36) | NA | 441.84 (109.46) | 412.29 (80.96) |
| Median (min, max) | 396.41 (209.95, 961.63) | 388.39 (209.95, 538.38) | NA | 421.27 (234.35, 961.63) | 391.09 (317.61, 562.02) |
| Missing | 651 | 428 | 100 | 66 | 57 |
| **Aβ40 tertiles**, n (%) |  |  |  |  |  |
| < 374.886 | 69 (33%) | 47 (38%) | NA | 19 (25%) | 3 (30%) |
| 374.886 - 422.785 | 71 (34%) | 46 (37%) | NA | 20 (26%) | 5 (50%) |
| > 422.785 | 69 (33%) | 30 (24%) | NA | 37 (49%) | 2 (20%) |
| **Aβ42**, mean (SD) | 43.91 (8.59) | 42.84 (7.33) | NA | 45.76 (10.27) | 42.94 (7.13) |
| Median (min, max) | 42.66 (24.67, 89.62) | 42.58 (26.96, 68.55) | NA | 42.64 (24.67, 89.62) | 43.07 (31.70, 53.16) |
| Missing | 651 | 428 | 100 | 66 | 57 |
| **Aβ42 tertiles**, n (%) |  |  |  |  |  |
| < 40.219 | 69 (33%) | 42 (34%) | NA | 24 (32%) | 3 (30%) |
| 40.219 - 45.870 | 71 (34%) | 45 (37%) | NA | 22 (29%) | 4 (40%) |
| > 45.870 | 69 (33%) | 36 (29%) | NA | 30 (39%) | 3 (30%) |
| **Aβ42/Aβ40**, mean (SD) | 0.11 (0.02) | 0.11 (0.02) | NA | 0.10 (0.01) | 0.10 (0.01) |
| Median (min, max) | 0.11 (0.08, 0.20) | 0.11 (0.09, 0.20) | NA | 0.10 (0.08, 0.19) | 0.11 (0.09, 0.12) |
| Missing | 651 | 428 | 100 | 66 | 57 |
| **Aβ42/Aβ40 tertiles**, n (%) |  |  |  |  |  |
| < 0.1040 | 69 (33%) | 32 (26%) | NA | 33 (43%) | 4 (40%) |
| 0.1040 - 0.1106 | 71 (34%) | 39 (32%) | NA | 29 (38%) | 3 (30%) |
| > 0.1106 | 69 (33%) | 52 (42%) | NA | 14 (18%) | 3 (30%) |

#### Supplemental Figure 1: Participants Followed at Each Yearly Interval

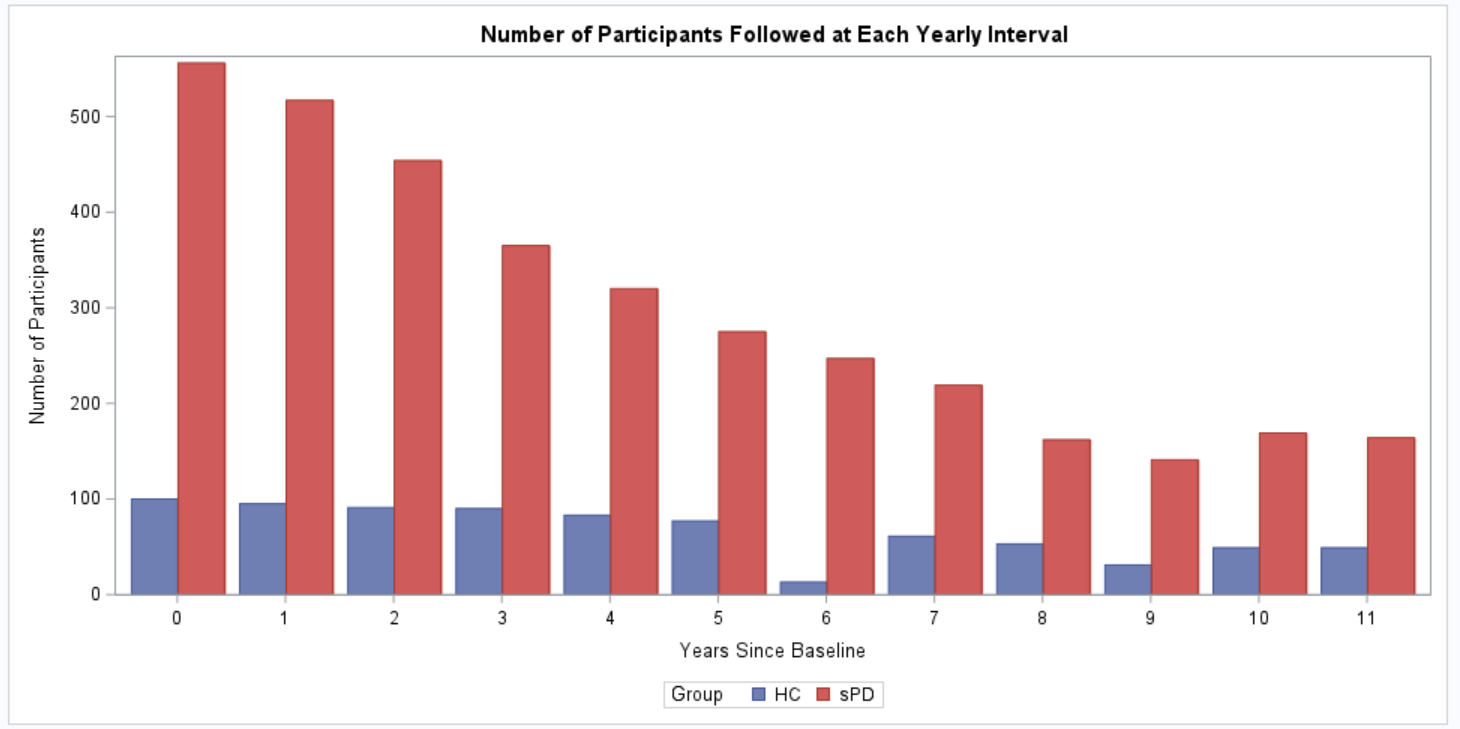

**Supplemental Figure 1: Participants Followed at Each Yearly Interval.** Number of participants followed at each yearly interval over 11 years of follow up. Individuals are not withdrawn for missing visits. Baseline, represented as 0 on the X axis, indicates study entry. HC = Healthy control cohort; PD = Sporadic Parkinson’s disease cohort.

#### Supplemental Figure 2: Box-Whisker Plot for Sporadic PD and Healthy Control at baseline

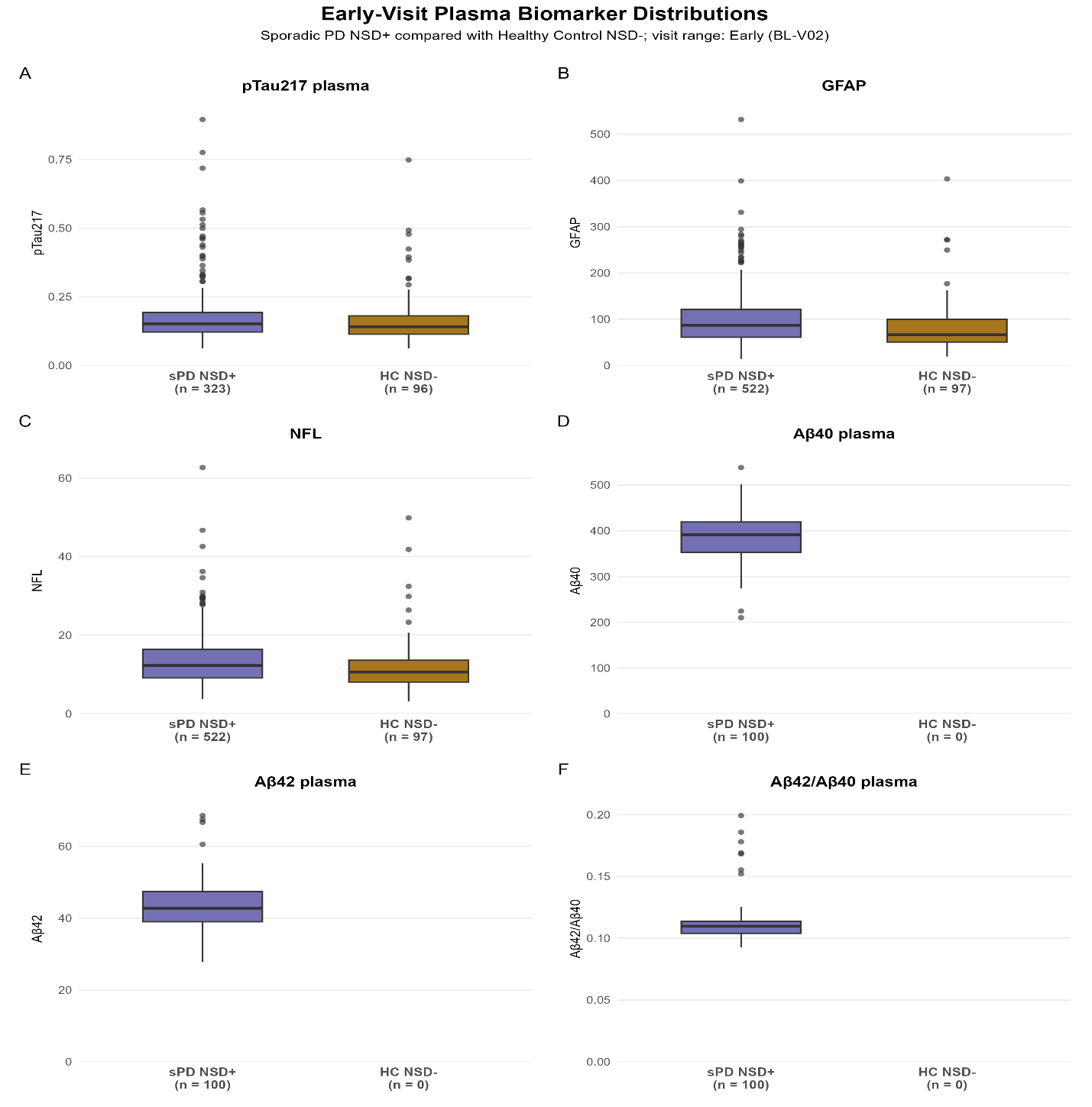

Supplemental Figure 2: Box-Whisker Plot for Sporadic PD and Healthy Control at baseline:

Boxes show the interquartile range (IQR), with the median indicated by the line inside each box. Whiskers extend to the most extreme observations within 1.5 × IQR of the box boundaries; values beyond the whiskers are shown as outliers. N denotes the number of participants with both a non-missing biomarker value and a corresponding non-missing assay date.

#### Supplemental Table 2a. Summary of NSD+ sPD participants by p-tau217 level.

|  | **pTau217 plasma tertiles** | | |  | | | | | |
| --- | --- | --- | --- | --- | --- | --- | --- | --- | --- |
| **Variable** | **<0.131** (N = 109) | **0.131-0.174** (N = 112) | **>0.174** (N = 110) | **Overall P-value** | **T1 vs. T2 P-value** | **T1 vs. T3 P-value** | **T2 vs. T3 P-value** | **Beta / OR (95% CI)** | **Continuous biomarker P-value** |
| **Sex**, n (%) |  |  |  | **0.0395** | **0.0154** | 0.6390 | **0.0497** |  |  |
| Male | 64 (59%) | 83 (74%) | 68 (62%) |  |  |  |  |  |  |
| Female | 45 (41%) | 29 (26%) | 42 (38%) |  |  |  |  |  |  |
| **Age (years)**, mean (SD) | 58.5 (8.4) | 61.7 (9.8) | 65.0 (9.2) | **<0.0001** | **0.0105** | **<0.0001** | **0.0088** |  |  |
| Median (min, max) | 59.1 (33.7, 77.0) | 62.5 (38.5, 80.2) | 67.0 (34.8, 84.9) |  |  |  |  |  |  |
| **Duration (years)**, mean (SD) | 0.6 (0.7) | 0.5 (0.4) | 0.6 (0.6) | 0.5033 |  |  |  |  |  |
| Median (min, max) | 0.4 (0.0, 3.0) | 0.3 (0.1, 1.9) | 0.4 (0.1, 2.7) |  |  |  |  |  |  |
| **Race**, n (%) |  |  |  | 0.3625 |  |  |  |  |  |
| White | 99 (92%) | 107 (96%) | 100 (91%) |  |  |  |  |  |  |
| Black/African American | 3 (3%) | 1 (1%) | 2 (2%) |  |  |  |  |  |  |
| Asian | 1 (1%) | 2 (2%) | 4 (4%) |  |  |  |  |  |  |
| Other | 5 (5%) | 2 (2%) | 4 (4%) |  |  |  |  |  |  |
| Missing | 1 | 0 | 0 |  |  |  |  |  |  |
| **Ethnicity**, n (%) |  |  |  | 0.9007 |  |  |  |  |  |
| Not Hispanic/Latino | 107 (98%) | 110 (98%) | 107 (97%) |  |  |  |  |  |  |
| Hispanic/Latino | 2 (2%) | 2 (2%) | 3 (3%) |  |  |  |  |  |  |
| **UPSIT Raw Score**, mean (SD) | 22.7 (7.7) | 20.7 (7.2) | 20.0 (7.5) | 0.6138^b^ |  |  |  | -2.1257(-9.9157,5.6644) | 0.5917^b^ |
| Median (min, max) | 23.0 (6.0, 37.0) | 20.0 (1.0, 38.0) | 19.0 (5.0, 36.0) |  |  |  |  |  |  |
| **Revised UPSIT percentile**, mean (SD) | 11.3 (15.5) | 9.4 (11.8) | 10.7 (13.5) | 0.9109 |  |  |  | 7.1323(-7.8246,22.0892) | 0.3489 |
| Median (min, max) | 5.5 (1.0, 78.5) | 5.0 (1.0, 91.5) | 5.5 (1.0, 64.0) |  |  |  |  |  |  |
| **Revised UPSIT percentile ≤ 15**, n (%) | 86 (79%) | 90 (80%) | 90 (82%) | 0.8626 |  |  |  | 1.6407(0.1220,22.0678) | 0.7089 |
| **NSD+**, n (%) |  |  |  | 0.1361^a^ |  |  |  | 8.1547(0.2693,>99.9999) | 0.2278^a^ |
| 2a | 1 (1%) | 2 (2%) | 3 (3%) |  |  |  |  |  |  |
| 2b | 26 (24%) | 31 (28%) | 16 (15%) |  |  |  |  |  |  |
| 3 | 74 (68%) | 68 (61%) | 78 (71%) |  |  |  |  |  |  |
| 4 | 8 (7%) | 11 (10%) | 13 (12%) |  |  |  |  |  |  |
| **MDS-UPDRS item 1.1**, n (%) |  |  |  | 0.4820^a^ |  |  |  | 0.2970(0.0269,3.2787) | 0.3218^a^ |
| Normal | 86 (79%) | 86 (77%) | 76 (69%) |  |  |  |  |  |  |
| Slight | 21 (19%) | 23 (21%) | 29 (26%) |  |  |  |  |  |  |
| Mild | 2 (2%) | 3 (3%) | 5 (5%) |  |  |  |  |  |  |
| **MDS-UPDRS part I**, mean (SD) | 4.8 (3.3) | 5.3 (4.1) | 6.1 (4.1) | 0.0624^f^ |  |  |  | 61.1309(-46.2802,168.5419) | 0.2637^f^ |
| Median (min, max) | 4 (0, 17) | 5 (0, 18) | 6 (0, 23) |  |  |  |  |  |  |
| **MDS-UPDRS part II**, mean (SD) | 5.4 (4.0) | 5.3 (4.2) | 6.6 (4.0) | **0.0107^f^** | 0.6094^f^ | **0.0224^f^** | **0.0043^f^** | 117.0262(9.2972,224.7551) | **0.0333^f^** |
| Median (min, max) | 5 (0, 22) | 4 (0, 20) | 6 (1, 18) |  |  |  |  |  |  |
| **MDS-UPDRS part III**, mean (SD) | 20.6 (8.5) | 20.7 (9.1) | 21.9 (9.0) | 0.7435^d^ |  |  |  | 0.2029(-0.8932,1.2990) | 0.7160^d^ |
| Median (min, max) | 20 (5, 46) | 19 (4, 41) | 21 (6, 51) |  |  |  |  |  |  |
| **MDS-UPDRS total score**, mean (SD) | 30.8 (11.9) | 31.4 (13.5) | 34.6 (13.7) | 0.2291^d^ |  |  |  | 0.7055(-0.6132,2.0242) | 0.2933^d^ |
| Median (min, max) | 29 (7, 70) | 30 (7, 68) | 36 (10, 70) |  |  |  |  |  |  |
| **MoCA total score**, mean (SD) | 27.3 (2.1) | 27.1 (2.5) | 27.0 (2.5) | 0.8434^f^ |  |  |  | 63.3852(-41.0386,167.8090) | 0.2333^f^ |
| Median (min, max) | 28 (22, 30) | 28 (17, 30) | 28 (17, 30) |  |  |  |  |  |  |
| **SCOPA-AUT total score**, mean (SD) | 8.7 (5.7) | 8.3 (5.8) | 11.4 (6.3) | **0.0024^f^** | 0.2251^f^ | **0.0290^f^** | **0.0006^f^** | 109.3100(3.7327,214.8873) | **0.0425^f^** |
| Median (min, max) | 8 (0, 39) | 7 (0, 32) | 10 (0, 32) |  |  |  |  |  |  |
| Missing | 1 | 2 | 2 |  |  |  |  |  |  |
| **% Age/sex-expected low putamen**, mean (SD) | 0.39 (0.13) | 0.41 (0.14) | 0.40 (0.15) | 0.3263 |  |  |  | 0.0610(-0.0935,0.2155) | 0.4377 |
| Median (min, max) | 0.36 (0.15, 0.99) | 0.40 (0.02, 0.95) | 0.38 (0.10, 0.87) |  |  |  |  |  |  |
| **Mean striatum CWM**, mean (SD) | 0.81 (0.21) | 0.80 (0.22) | 0.77 (0.21) | 0.5917^b^ |  |  |  | -0.0336(-0.2648,0.1977) | 0.7754^b^ |
| Median (min, max) | 0.80 (0.36, 1.40) | 0.79 (0.13, 1.36) | 0.76 (0.27, 1.29) |  |  |  |  |  |  |
| **Mean putamen CWM**, mean (SD) | 0.71 (0.21) | 0.74 (0.23) | 0.72 (0.23) | 0.5231^d^ |  |  |  | -0.0308(-0.1811,0.1194) | 0.6864^d^ |
| Median (min, max) | 0.69 (0.32, 1.55) | 0.72 (0.10, 1.56) | 0.68 (0.19, 1.27) |  |  |  |  |  |  |
| **Cognitive categorization**, n (%) |  |  |  | 0.3568 ^a^ |  |  |  | <0.0001(<0.0001,39.8169) | 0.0952 ^a^ |
| Normal cognition | 20 (91%) | 24 (86%) | 23 (96%) |  |  |  |  |  |  |
| Mild cognitive impairment (MCI) | 2 (9%) | 4 (14%) | 1 (4%) |  |  |  |  |  |  |
| Missing | 87 | 84 | 86 |  |  |  |  |  |  |

#### Supplemental Table 2b. Summary of HC participants by p-tau217 level.

|  | **pTau217 plasma tertiles** | | |  | | | | | |
| --- | --- | --- | --- | --- | --- | --- | --- | --- | --- |
| **Variable** | **<0.131** (N = 40) | **0.131-0.174** (N = 30) | **>0.174** (N = 27) | **Overall P-value** | **T1 vs. T2 P-value** | **T1 vs. T3 P-value** | **T2 vs. T3 P-value** | **Beta / OR (95% CI)** | **Continuous biomarker P-value** |
| **Sex**, n (%) |  |  |  | 0.5806 |  |  |  |  |  |
| Male | 23 (58%) | 16 (53%) | 18 (67%) |  |  |  |  |  |  |
| Female | 17 (43%) | 14 (47%) | 9 (33%) |  |  |  |  |  |  |
| **Age (years)**, mean (SD) | 55.0 (12.6) | 59.4 (11.4) | 64.1 (11.4) | **0.0209** | 0.2063 | **0.0069** | 0.1154 |  |  |
| Median (min, max) | 57.1 (30.6, 78.8) | 61.1 (31.9, 76.8) | 64.2 (32.3, 81.3) |  |  |  |  |  |  |
| Missing | 40 | 30 | 27 |  |  |  |  |  |  |
| **Race**, n (%) |  |  |  | 0.8268 |  |  |  |  |  |
| White | 36 (90%) | 28 (93%) | 24 (89%) |  |  |  |  |  |  |
| Black/African American | 3 (8%) | 1 (3%) | 2 (7%) |  |  |  |  |  |  |
| Asian | 0 | 1 (3%) | 0 |  |  |  |  |  |  |
| Other | 1 (3%) | 0 | 1 (4%) |  |  |  |  |  |  |
| **Ethnicity**, n (%) |  |  |  | 1.0000 |  |  |  |  |  |
| Not Hispanic/Latino | 39 (98%) | 29 (97%) | 27 (100%) |  |  |  |  |  |  |
| Hispanic/Latino | 1 (3%) | 1 (3%) | 0 |  |  |  |  |  |  |
| **UPSIT Raw Score**, mean (SD) | 34.4 (3.7) | 34.2 (4.0) | 34.1 (3.4) | 0.8160^f^ |  |  |  | 19.5948(-37.4579,76.6476) | 0.4969 ^f^ |
| Median (min, max) | 35.0 (21.0, 40.0) | 35.5 (21.0, 39.0) | 34.0 (26.0, 39.0) |  |  |  |  |  |  |
| **Revised UPSIT percentile**, mean (SD) | 56.6 (30.3) | 62.8 (28.4) | 61.6 (26.3) | 0.6085 |  |  |  | 29.6456(-27.6285,86.9196) | 0.3067 |
| Median (min, max) | 61.3 (2.0, 100.0) | 66.5 (9.0, 97.5) | 59.5 (14.5, 98.5) |  |  |  |  |  |  |
| **Revised UPSIT percentile ≤ 15**, n (%) | 5 (13%) | 3 (10%) | 2 (7%) | 0.9137 |  |  |  | 90933.57(0.0093,8.89E11) | 0.1644 |
| **MDS-UPDRS item 1.1**, n (%) |  |  |  | 0.9405 ^a^ |  |  |  | 6.2664(<0.0001,>99.9999) | 0.7584 ^a^ |
| Normal | 37 (93%) | 29 (100%) | 25 (93%) |  |  |  |  |  |  |
| Slight | 2 (5%) | 0 | 2 (7%) |  |  |  |  |  |  |
| Mild | 1 (3%) | 0 | 0 |  |  |  |  |  |  |
| Missing | 0 | 1 | 0 |  |  |  |  |  |  |
| **MDS-UPDRS part I**, mean (SD) | 3.3 (3.1) | 1.8 (2.5) | 2.7 (2.5) | 0.1091 ^c^ |  |  |  | -0.6521(-2.3095,1.0053) | 0.4345 ^c^ |
| Median (min, max) | 3 (0, 12) | 1 (0, 11) | 2 (0, 8) |  |  |  |  |  |  |
| Missing | 0 | 1 | 0 |  |  |  |  |  |  |
| **MDS-UPDRS part II**, mean (SD) | 0.6 (1.2) | 0.3 (0.8) | 0.7 (1.3) | 0.4301 ^f^ |  |  |  | 4.8765(-41.8353,51.5883) | 0.8362 ^f^ |
| Median (min, max) | 0 (0, 6) | 0 (0, 3) | 0 (0, 5) |  |  |  |  |  |  |
| Missing | 0 | 1 | 0 |  |  |  |  |  |  |
| **MDS-UPDRS part III**, mean (SD) | 0.6 (1.0) | 0.7 (1.3) | 2.0 (3.4) | 0.0682 ^c^ |  |  |  | 2.9034(0.6210,5.1857) | **0.0145 ^c^** |
| Median (min, max) | 0 (0, 4) | 0 (0, 5) | 0 (0, 13) |  |  |  |  |  |  |
| Missing | 0 | 1 | 0 |  |  |  |  |  |  |
| **MDS-UPDRS total score**, mean (SD) | 4.4 (3.9) | 2.9 (3.5) | 5.4 (5.2) | 0.0607 ^c^ |  |  |  | 0.9461(-0.8470,2.7392) | 0.2961 ^c^ |
| Median (min, max) | 4 (0, 15) | 2 (0, 13) | 4 (0, 17) |  |  |  |  |  |  |
| Missing | 0 | 1 | 0 |  |  |  |  |  |  |
| **MoCA total score**, mean (SD) | 28.6 (1.2) | 28.5 (1.0) | 27.9 (1.1) | 0.1397 ^f^ |  |  |  | -63.1743(-119.3231,-7.0255) | **0.0279 ^f^** |
| Median (min, max) | 29 (27, 30) | 29 (27, 30) | 28 (27, 30) |  |  |  |  |  |  |
| **SCOPA-AUT total score**, mean (SD) | 6.0 (4.5) | 5.4 (3.0) | 5.6 (3.1) | 0.2310 ^d^ |  |  |  | -1.1867(-2.6775,0.3041) | 0.1173 ^d^ |
| Median (min, max) | 5 (0, 18) | 5 (0, 12) | 5 (1, 14) |  |  |  |  |  |  |
| **% Age/sex-expected low putamen**, mean (SD) | 1.00 (0.13) | 1.05 (0.16) | 0.94 (0.14) | **0.0242** | 0.1198 | 0.1126 | **0.0101** | -0.0596(-0.3595,0.2403) | 0.6939 |
| Median (min, max) | 1.00 (0.75, 1.30) | 1.05 (0.70, 1.35) | 0.93 (0.65, 1.23) |  |  |  |  |  |  |
| Missing | 1 | 0 | 1 |  |  |  |  |  |  |
| **Mean striatum CWM**, mean (SD) | 1.47 (0.21) | 1.50 (0.31) | 1.29 (0.18) | **0.0359 ^b^** | 0.1610 ^b^ | 0.1752 ^b^ | **0.0103 ^b^** | -0.0246(-0.4723,0.4231) | 0.9134 ^b^ |
| Median (min, max) | 1.44 (1.08, 1.85) | 1.44 (0.96, 2.17) | 1.27 (1.01, 1.65) |  |  |  |  |  |  |
| Missing | 1 | 0 | 1 |  |  |  |  |  |  |
| **Mean putamen CWM**, mean (SD) | 1.59 (0.20) | 1.65 (0.31) | 1.44 (0.21) | **0.0275 ^b^** | 0.1925 ^b^ | 0.1187 ^b^ | **0.0075 ^b^** | -0.0931(-0.5956,0.4095) | 0.7138 ^b^ |
| Median (min, max) | 1.59 (1.17, 1.98) | 1.62 (0.99, 2.22) | 1.44 (1.00, 1.82) |  |  |  |  |  |  |
| Missing | 1 | 0 | 1 |  |  |  |  |  |  |
| **Cognitive categorization**, n (%) |  |  |  |  |  |  |  |  |  |
| Normal cognition | 6 (100%) | 2 (100%) | 7 (100%) |  |  |  |  |  |  |
| Missing | 34 | 28 | 20 |  |  |  |  |  |  |

#### Supplemental Table 2c. Summary of sPD participants by NFL level.

|  | **NFL tertiles** | | |  | | | | | |
| --- | --- | --- | --- | --- | --- | --- | --- | --- | --- |
| **Variable** | **<9.877088349** (N = 172) | **9.877088349-14.53844902** (N = 163) | **>14.53844902** (N = 187) | **Overall P-value** | **T1 vs. T2 P-value** | **T1 vs. T3 P-value** | **T2 vs. T3 P-value** | **Beta / OR (95% CI)** | **Continuous biomarker P-value** |
| **Sex**, n (%) |  |  |  | 0.8562 |  |  |  |  |  |
| Male | 109 (63%) | 99 (61%) | 114 (61%) |  |  |  |  |  |  |
| Female | 63 (37%) | 64 (39%) | 73 (39%) |  |  |  |  |  |  |
| **Age (years)**, mean (SD) | 55.6 (8.3) | 62.9 (7.1) | 69.5 (6.6) | **<0.0001** | **<0.0001** | **<0.0001** | **<0.0001** |  |  |
| Median (min, max) | 55.9 (33.7, 74.1) | 63.5 (42.7, 78.0) | 70.6 (48.5, 84.9) |  |  |  |  |  |  |
| **Duration (years)**, mean (SD) | 0.7 (0.7) | 0.6 (0.5) | 0.7 (0.5) | 0.2964 |  |  |  |  |  |
| Median (min, max) | 0.5 (0.0, 3.2) | 0.4 (0.1, 3.0) | 0.5 (0.1, 2.9) |  |  |  |  |  |  |
| **Race**, n (%) |  |  |  | 0.6373 |  |  |  |  |  |
| White | 156 (92%) | 153 (94%) | 175 (94%) |  |  |  |  |  |  |
| Black/African American | 2 (1%) | 5 (3%) | 1 (1%) |  |  |  |  |  |  |
| Asian | 4 (2%) | 1 (1%) | 5 (3%) |  |  |  |  |  |  |
| Other | 8 (5%) | 4 (2%) | 5 (3%) |  |  |  |  |  |  |
| Missing | 2 | 0 | 1 |  |  |  |  |  |  |
| **Ethnicity**, n (%) |  |  |  | 0.6462 |  |  |  |  |  |
| Not Hispanic/Latino | 167 (97%) | 154 (96%) | 182 (97%) |  |  |  |  |  |  |
| Hispanic/Latino | 5 (3%) | 7 (4%) | 5 (3%) |  |  |  |  |  |  |
| Missing | 0 | 2 | 0 |  |  |  |  |  |  |
| **UPSIT Raw Score**, mean (SD) | 24.0 (6.9) | 21.3 (7.2) | 19.2 (7.2) | **0.0129 ^f^** | **0.0424 ^f^** | **0.0034 ^f^** | 0.1749 ^f^ | -1.9238(-4.3193,0.4716) | 0.1152 ^f^ |
| Median (min, max) | 24.0 (7.0, 38.0) | 21.0 (1.0, 37.0) | 18.0 (5.0, 36.0) |  |  |  |  |  |  |
| **Revised UPSIT percentile**, mean (SD) | 11.9 (15.8) | 10.7 (12.2) | 10.2 (11.6) | 0.8510 |  |  |  | -0.0937(-0.2807,0.0934) | 0.3256 |
| Median (min, max) | 6.0 (1.0, 91.5) | 7.0 (1.0, 69.5) | 6.5 (1.0, 64.0) |  |  |  |  |  |  |
| **Revised UPSIT percentile ≤ 15**, n (%) | 129 (76%) | 129 (80%) | 144 (80%) | 0.6575 |  |  |  | 0.9929(0.9586,1.0284) | 0.6909 |
| Missing | 3 | 2 | 6 |  |  |  |  |  |  |
| **NSD+**, n (%) |  |  |  | 0.7355 ^a^ |  |  |  | 0.9748(0.9324,1.0191) | 0.2596 ^a^ |
| 2a | 1 (1%) | 3 (2%) | 4 (2%) |  |  |  |  |  |  |
| 2b | 28 (16%) | 25 (15%) | 21 (11%) |  |  |  |  |  |  |
| 3 | 130 (76%) | 130 (80%) | 149 (80%) |  |  |  |  |  |  |
| 4 | 13 (8%) | 5 (3%) | 13 (7%) |  |  |  |  |  |  |
| **MDS-UPDRS item 1.1**, n (%) |  |  |  | 0.6635 ^a^ |  |  |  | 1.0242(0.9837,1.0664) | 0.2455 ^a^ |
| Normal | 136 (79%) | 118 (72%) | 132 (71%) |  |  |  |  |  |  |
| Slight | 31 (18%) | 41 (25%) | 53 (28%) |  |  |  |  |  |  |
| Mild | 5 (3%) | 4 (2%) | 2 (1%) |  |  |  |  |  |  |
| **MDS-UPDRS part I**, mean (SD) | 5.4 (3.7) | 5.4 (3.4) | 6.0 (3.7) | 0.8121 ^f^ |  |  |  | -0.5978(-3.1177,1.9221) | 0.6414 ^f^ |
| Median (min, max) | 5 (0, 17) | 5 (0, 18) | 5 (0, 23) |  |  |  |  |  |  |
| **MDS-UPDRS part II**, mean (SD) | 6.0 (3.9) | 5.4 (3.2) | 6.6 (3.8) | 0.0538 ^d^ |  |  |  | 0.0025(-0.0103,0.0154) | 0.6965 ^d^ |
| Median (min, max) | 5 (0, 22) | 5 (0, 18) | 6 (1, 18) |  |  |  |  |  |  |
| **MDS-UPDRS part III**, mean (SD) | 21.1 (9.1) | 20.7 (8.5) | 24.8 (9.7) | **0.0165 ^d^** | 0.1979 ^d^ | 0.1961 ^d^ | **0.0045 ^d^** | 0.0022(-0.0144,0.0189) | 0.7928 ^d^ |
| Median (min, max) | 21 (4, 46) | 20 (5, 46) | 23 (6, 52) |  |  |  |  |  |  |
| **MDS-UPDRS total score**, mean (SD) | 32.5 (12.6) | 31.5 (11.7) | 37.3 (13.0) | **0.0087 ^d^** | 0.0879 ^d^ | 0.2942 ^d^ | **0.0028 ^d^** | 0.0042(-0.0143,0.0228) | 0.6551 ^d^ |
| Median (min, max) | 32 (8, 70) | 30 (7, 70) | 37 (11, 68) |  |  |  |  |  |  |
| **MoCA total score**, mean (SD) | 27.6 (1.9) | 27.0 (2.5) | 26.8 (2.5) | 0.8993 ^f^ |  |  |  | 0.4842(-1.9752,2.9436) | 0.6991 ^f^ |
| Median (min, max) | 28 (20, 30) | 28 (17, 30) | 27 (17, 30) |  |  |  |  |  |  |
| **SCOPA-AUT total score**, mean (SD) | 8.7 (5.6) | 10.3 (6.3) | 9.9 (5.1) | 0.5041 ^f^ |  |  |  | 0.4004(-2.0900,2.8908) | 0.7522 ^f^ |
| Median (min, max) | 8 (0, 39) | 10 (0, 32) | 10 (0, 26) |  |  |  |  |  |  |
| Missing | 1 | 3 | 2 |  |  |  |  |  |  |
| **% Age/sex-expected low putamen**, mean (SD) | 0.38 (0.12) | 0.42 (0.14) | 0.43 (0.15) | **0.0036** | **0.0262** | **0.0010** | 0.3362 | 0.0031(0.0012,0.0050) | **0.0015** |
| Median (min, max) | 0.37 (0.15, 0.99) | 0.39 (0.02, 0.95) | 0.43 (0.10, 0.87) |  |  |  |  |  |  |
| **Mean striatum CWM**, mean (SD) | 0.83 (0.20) | 0.81 (0.22) | 0.75 (0.20) | 0.6013 ^b^ |  |  |  | -0.0007(-0.0042,0.0027) | 0.6708 ^b^ |
| Median (min, max) | 0.82 (0.32, 1.40) | 0.78 (0.13, 1.33) | 0.75 (0.22, 1.29) |  |  |  |  |  |  |
| **Mean putamen CWM**, mean (SD) | 0.72 (0.20) | 0.75 (0.24) | 0.74 (0.23) | 0.6819 ^d^ |  |  |  | 0.0006(-0.0017,0.0028) | 0.6161 ^d^ |
| Median (min, max) | 0.70 (0.28, 1.55) | 0.71 (0.10, 1.56) | 0.71 (0.19, 1.25) |  |  |  |  |  |  |
| **Cognitive categorization**, n (%) |  |  |  | 0.7922 ^a^ |  |  |  | 0.9877(0.9010,1.0827) | 0.7916 ^a^ |
| Normal cognition | 77 (97%) | 73 (94%) | 94 (89%) |  |  |  |  |  |  |
| Mild cognitive impairment (MCI) | 2 (3%) | 5 (6%) | 12 (11%) |  |  |  |  |  |  |
| Missing | 93 | 85 | 81 |  |  |  |  |  |  |

#### Supplemental Table 2d. Summary of HC participants by NFL level.

|  | **NFL tertiles** | | |  | | | | | |
| --- | --- | --- | --- | --- | --- | --- | --- | --- | --- |
| **Variable** | **<9.877088349** (N = 44) | **9.877088349-14.53844902** (N = 33) | **>14.53844902** (N = 20) | **Overall P-value** | **T1 vs. T2 P-value** | **T1 vs. T3 P-value** | **T2 vs. T3 P-value** | **Beta / OR (95% CI)** | **Continuous biomarker P-value** |
| **Sex**, n (%) |  |  |  | 0.5245 |  |  |  |  |  |
| Male | 24 (55%) | 22 (67%) | 11 (55%) |  |  |  |  |  |  |
| Female | 20 (45%) | 11 (33%) | 9 (45%) |  |  |  |  |  |  |
| **Age (years)**, mean (SD) | 49.7 (10.0) | 64.4 (6.5) | 70.0 (10.0) | **<0.0001** | **<0.0001** | **<0.0001** | **0.0089** |  |  |
| Median (min, max) | 53.3 (30.6, 66.8) | 64.8 (50.3, 76.6) | 72.8 (43.2, 81.3) |  |  |  |  |  |  |
| Missing | 44 | 33 | 20 |  |  |  |  |  |  |
| **Race**, n (%) |  |  |  | 0.8186 |  |  |  |  |  |
| White | 40 (91%) | 29 (88%) | 19 (95%) |  |  |  |  |  |  |
| Black/African American | 4 (9%) | 1 (3%) | 1 (5%) |  |  |  |  |  |  |
| Asian | 0 | 1 (3%) | 0 |  |  |  |  |  |  |
| Other | 0 | 2 (6%) | 0 |  |  |  |  |  |  |
| **Ethnicity**, n (%) |  |  |  | 1.0000 |  |  |  |  |  |
| Not Hispanic/Latino | 43 (98%) | 32 (97%) | 20 (100%) |  |  |  |  |  |  |
| Hispanic/Latino | 1 (2%) | 1 (3%) | 0 |  |  |  |  |  |  |
| **UPSIT Raw Score**, mean (SD) | 35.7 (2.8) | 31.7 (3.8) | 35.3 (3.0) | **<0.0001 ^f^** | **0.0003 ^f^** | 0.5117^f^ | **<0.0001 ^f^** | 0.6114(-0.3073,1.5301) | 0.1896 ^f^ |
| Median (min, max) | 36.0 (27.0, 39.0) | 33.0 (21.0, 37.0) | 36.0 (28.0, 40.0) |  |  |  |  |  |  |
| **Revised UPSIT percentile**, mean (SD) | 66.1 (27.7) | 42.5 (23.0) | 75.1 (25.1) | **<0.0001** | **0.0001** | 0.1066 | **<0.0001** | 0.2397(-0.5715,1.0508) | 0.5589 |
| Median (min, max) | 74.0 (2.0, 97.5) | 40.0 (6.0, 88.5) | 85.5 (10.0, 100.0) |  |  |  |  |  |  |
| **Revised UPSIT percentile ≤ 15**, n (%) | 3 (7%) | 6 (18%) | 1 (5%) | 0.2837 |  |  |  | 0.9828(0.9063,1.0657) | 0.6750 |
| **MDS-UPDRS item 1.1**, n (%) |  |  |  | 0.9017 ^a^ |  |  |  | 0.9379(0.8334,1.0556) | 0.2879 ^a^ |
| Normal | 41 (93%) | 32 (100%) | 18 (90%) |  |  |  |  |  |  |
| Slight | 2 (5%) | 0 | 2 (10%) |  |  |  |  |  |  |
| Mild | 1 (2%) | 0 | 0 |  |  |  |  |  |  |
| Missing | 0 | 1 | 0 |  |  |  |  |  |  |
| **MDS-UPDRS part I**, mean (SD) | 2.4 (2.6) | 2.4 (2.4) | 3.7 (3.8) | 0.6277 ^c^ |  |  |  | 0.0091(-0.0154,0.0337) | 0.4607 ^c^ |
| Median (min, max) | 2 (0, 10) | 3 (0, 8) | 2 (0, 12) |  |  |  |  |  |  |
| Missing | 0 | 1 | 0 |  |  |  |  |  |  |
| **MDS-UPDRS part II**, mean (SD) | 0.2 (0.5) | 0.8 (1.3) | 0.9 (1.5) | 0.3004 ^c^ |  |  |  | 0.0240(-0.0013,0.0493) | 0.0619 ^c^ |
| Median (min, max) | 0 (0, 2) | 0 (0, 6) | 0 (0, 5) |  |  |  |  |  |  |
| Missing | 0 | 1 | 0 |  |  |  |  |  |  |
| **MDS-UPDRS part III**, mean (SD) | 0.5 (0.9) | 0.8 (1.7) | 2.5 (3.6) | 0.9452 ^c^ |  |  |  | 0.0402(-0.0254,0.1058) | 0.2203 ^c^ |
| Median (min, max) | 0 (0, 3) | 0 (0, 7) | 1 (0, 13) |  |  |  |  |  |  |
| Missing | 0 | 1 | 0 |  |  |  |  |  |  |
| **MDS-UPDRS total score**, mean (SD) | 3.1 (3.1) | 4.0 (3.9) | 7.0 (5.7) | 0.4782 ^c^ |  |  |  | 0.0224(-0.0039,0.0487) | 0.0934 ^c^ |
| Median (min, max) | 2 (0, 12) | 3 (0, 15) | 7 (0, 17) |  |  |  |  |  |  |
| Missing | 0 | 1 | 0 |  |  |  |  |  |  |
| **MoCA total score**, mean (SD) | 28.8 (1.2) | 28.0 (1.1) | 28.1 (1.0) | 0.1201 ^f^ |  |  |  | -0.4502(-1.3742,0.4737) | 0.3357 ^f^ |
| Median (min, max) | 29 (27, 30) | 28 (27, 30) | 28 (27, 30) |  |  |  |  |  |  |
| **SCOPA-AUT total score**, mean (SD) | 4.4 (2.9) | 6.2 (3.8) | 7.8 (4.0) | 0.7882 ^d^ |  |  |  | 0.0257(0.0021,0.0494) | **0.0334 ^d^** |
| Median (min, max) | 4 (0, 12) | 6 (0, 17) | 8 (1, 18) |  |  |  |  |  |  |
| **% Age/sex-expected low putamen**, mean (SD) | 1.00 (0.14) | 0.95 (0.12) | 1.07 (0.16) | **0.0160** | 0.0711 | 0.1418 | **0.0059** | 0.0046(0.0005,0.0087) | **0.0284** |
| Median (min, max) | 1.00 (0.65, 1.30) | 0.93 (0.70, 1.30) | 1.05 (0.68, 1.35) |  |  |  |  |  |  |
| Missing | 1 | 1 | 0 |  |  |  |  |  |  |
| **Mean striatum CWM**, mean (SD) | 1.54 (0.23) | 1.29 (0.17) | 1.43 (0.30) | **0.0049 ^b^** | 0.0974 ^b^ | 0.1967 ^b^ | **0.0018^b^** | 0.0051(0.0004,0.0099) | **0.0350 ^c^** |
| Median (min, max) | 1.51 (1.02, 2.17) | 1.27 (0.96, 1.85) | 1.37 (1.01, 2.01) |  |  |  |  |  |  |
| Missing | 1 | 1 | 0 |  |  |  |  |  |  |
| **Mean putamen CWM**, mean (SD) | 1.61 (0.24) | 1.46 (0.21) | 1.64 (0.29) | **0.0139 ^b^** | 0.2538 ^b^ | 0.1366 ^b^ | **0.0039^b^** | 0.0109(0.0032,0.0186) | **0.0061 ^b^** |
| Median (min, max) | 1.61 (1.00, 2.22) | 1.45 (0.99, 1.98) | 1.62 (1.17, 2.15) |  |  |  |  |  |  |
| Missing | 1 | 1 | 0 |  |  |  |  |  |  |
| **Cognitive categorization**, n (%) |  |  |  |  |  |  |  |  |  |
| Normal cognition | 7 (100%) | 5 (100%) | 3 (100%) |  |  |  |  |  |  |
| Missing | 37 | 28 | 17 |  |  |  |  |  |  |

#### Supplemental Table 2e. Summary of sPD participants by GFAP level.

|  | **GFAP tertiles** | | |  | | | | | |
| --- | --- | --- | --- | --- | --- | --- | --- | --- | --- |
| **Variable** | **<64.47757262** (N = 144) | **64.47757262-105.3411683** (N = 191) | **>105.3411683** (N = 187) | **Overall P-value** | **T1 vs. T2 P-value** | **T1 vs. T3 P-value** | **T2 vs. T3 P-value** | **Beta / OR (95% CI)** | **Continuous biomarker P-value** |
| **Sex**, n (%) |  |  |  | **0.0139** | 0.2439 | **0.0044** | 0.0652 |  |  |
| Male | 100 (69%) | 121 (63%) | 101 (54%) |  |  |  |  |  |  |
| Female | 44 (31%) | 70 (37%) | 86 (46%) |  |  |  |  |  |  |
| **Age (years)**, mean (SD) | 55.8 (9.0) | 62.8 (7.8) | 68.5 (7.1) | **<0.0001** | **<0.0001** | **<0.0001** | **<0.0001** |  |  |
| Median (min, max) | 55.7 (33.7, 75.8) | 63.4 (39.2, 80.9) | 69.6 (46.6, 84.9) |  |  |  |  |  |  |
| **Duration (years)**, mean (SD) | 0.7 (0.6) | 0.7 (0.6) | 0.7 (0.6) | 0.7194 |  |  |  |  |  |
| Median (min, max) | 0.5 (0.0, 2.9) | 0.5 (0.1, 3.2) | 0.5 (0.1, 3.0) |  |  |  |  |  |  |
| **Race**, n (%) |  |  |  | 0.7890 |  |  |  |  |  |
| White | 132 (92%) | 178 (94%) | 174 (93%) |  |  |  |  |  |  |
| Black/African American | 2 (1%) | 2 (1%) | 4 (2%) |  |  |  |  |  |  |
| Asian | 4 (3%) | 3 (2%) | 3 (2%) |  |  |  |  |  |  |
| Other | 5 (3%) | 6 (3%) | 6 (3%) |  |  |  |  |  |  |
| Missing | 1 | 2 | 0 |  |  |  |  |  |  |
| **Ethnicity**, n (%) |  |  |  | 0.2243 |  |  |  |  |  |
| Not Hispanic/Latino | 139 (97%) | 180 (95%) | 184 (98%) |  |  |  |  |  |  |
| Hispanic/Latino | 5 (3%) | 9 (5%) | 3 (2%) |  |  |  |  |  |  |
| Missing | 0 | 2 | 0 |  |  |  |  |  |  |
| **UPSIT Raw Score**, mean (SD) | 23.5 (7.4) | 21.4 (7.4) | 19.9 (6.9) | 0.1840 ^f^ |  |  |  | -0.1446(-0.3979,0.1087) | 0.2627 ^f^ |
| Median (min, max) | 24.0 (7.0, 38.0) | 21.0 (1.0, 37.0) | 19.0 (5.0, 37.0) |  |  |  |  |  |  |
| **Revised UPSIT percentile**, mean (SD) | 11.5 (15.0) | 11.5 (14.2) | 9.9 (10.7) | 0.7173 |  |  |  | -0.0093(-0.0302,0.0116) | 0.3829 |
| Median (min, max) | 6.0 (1.0, 91.5) | 6.0 (1.0, 78.5) | 7.0 (1.0, 64.0) |  |  |  |  |  |  |
| **Revised UPSIT percentile ≤ 15**, n (%) | 107 (76%) | 146 (78%) | 149 (82%) | 0.3917 |  |  |  | 0.9978(0.9937,1.0020) | 0.3022 |
| Missing | 3 | 3 | 5 |  |  |  |  |  |  |
| **NSD+**, n (%) |  |  |  | 0.5363 ^a^ |  |  |  | 1.0010(0.9956,1.0064) | 0.7187 ^a^ |
| 2a | 1 (1%) | 2 (1%) | 5 (3%) |  |  |  |  |  |  |
| 2b | 26 (18%) | 24 (13%) | 24 (13%) |  |  |  |  |  |  |
| 3 | 105 (73%) | 159 (83%) | 145 (78%) |  |  |  |  |  |  |
| 4 | 12 (8%) | 6 (3%) | 13 (7%) |  |  |  |  |  |  |
| **MDS-UPDRS item 1.1**, n (%) |  |  |  | 0.9588 ^a^ |  |  |  | 1.0014(0.9972,1.0055) | 0.5154 ^a^ |
| Normal | 115 (80%) | 141 (74%) | 130 (70%) |  |  |  |  |  |  |
| Slight | 24 (17%) | 46 (24%) | 55 (29%) |  |  |  |  |  |  |
| Mild | 5 (3%) | 4 (2%) | 2 (1%) |  |  |  |  |  |  |
| **MDS-UPDRS part I**, mean (SD) | 5.3 (3.7) | 5.6 (3.4) | 6.0 (3.8) | 0.9361 ^f^ |  |  |  | 0.0138(-0.2537,0.2813) | 0.9193 ^f^ |
| Median (min, max) | 5 (0, 18) | 5 (0, 17) | 5 (0, 23) |  |  |  |  |  |  |
| **MDS-UPDRS part II**, mean (SD) | 5.9 (4.0) | 6.0 (3.6) | 6.0 (3.6) | 0.7120 ^d^ |  |  |  | <0.0001(-0.0013,0.0015) | 0.8974 ^d^ |
| Median (min, max) | 5 (0, 20) | 5 (0, 22) | 5 (1, 18) |  |  |  |  |  |  |
| **MDS-UPDRS part III**, mean (SD) | 21.2 (9.5) | 22.0 (8.7) | 23.5 (9.7) | 0.9125 ^d^ |  |  |  | 0.0011(-0.0007,0.0028) | 0.2319 ^d^ |
| Median (min, max) | 20 (4, 46) | 21 (5, 51) | 22 (6, 52) |  |  |  |  |  |  |
| **MDS-UPDRS total score**, mean (SD) | 32.4 (13.2) | 33.6 (11.8) | 35.5 (13.1) | 0.9596 ^d^ |  |  |  | 0.0010(-0.0009,0.0030) | 0.2967 ^d^ |
| Median (min, max) | 32 (7, 70) | 33 (7, 70) | 34 (11, 68) |  |  |  |  |  |  |
| **MoCA total score**, mean (SD) | 27.6 (2.1) | 27.0 (2.2) | 26.9 (2.5) | 0.3216 ^f^ |  |  |  | -0.0395(-0.3005,0.2215) | 0.7664 ^f^ |
| Median (min, max) | 28 (19, 30) | 27 (20, 30) | 27 (17, 30) |  |  |  |  |  |  |
| **SCOPA-AUT total score**, mean (SD) | 8.7 (6.2) | 9.8 (5.4) | 10.2 (5.5) | 0.4851 ^f^ |  |  |  | 0.0652(-0.1987,0.3291) | 0.6277 ^f^ |
| Median (min, max) | 7 (0, 39) | 10 (0, 27) | 10 (0, 28) |  |  |  |  |  |  |
| Missing | 1 | 2 | 3 |  |  |  |  |  |  |
| **% Age/sex-expected low putamen**, mean (SD) | 0.38 (0.12) | 0.42 (0.13) | 0.43 (0.16) | **0.0249** | **0.0171** | **0.0160** | 0.7754 | 0.0003(0.0000,0.0005) | **0.0234** |
| Median (min, max) | 0.37 (0.15, 0.99) | 0.40 (0.02, 0.80) | 0.40 (0.10, 0.95) |  |  |  |  |  |  |
| **Mean striatum CWM**, mean (SD) | 0.83 (0.21) | 0.81 (0.20) | 0.76 (0.22) | 0.5283 ^b^ |  |  |  | -0.0002(-0.0006,0.0001) | 0.2142 ^b^ |
| Median (min, max) | 0.81 (0.36, 1.40) | 0.79 (0.13, 1.33) | 0.77 (0.24, 1.27) |  |  |  |  |  |  |
| **Mean putamen CWM**, mean (SD) | 0.72 (0.21) | 0.74 (0.21) | 0.74 (0.25) | 0.6610 ^d^ |  |  |  | >-0.0001(-0.0003,0.0002) | 0.7662 ^d^ |
| Median (min, max) | 0.71 (0.28, 1.55) | 0.71 (0.10, 1.34) | 0.72 (0.19, 1.56) |  |  |  |  |  |  |
| **Cognitive categorization**, n (%) |  |  |  | 0.6884 ^a^ |  |  |  | 1.0004(0.9915,1.0094) | 0.9301 ^a^ |
| Normal cognition | 63 (97%) | 101 (94%) | 80 (89%) |  |  |  |  |  |  |
| Mild cognitive impairment (MCI) | 2 (3%) | 7 (6%) | 10 (11%) |  |  |  |  |  |  |
| Missing | 79 | 83 | 97 |  |  |  |  |  |  |

#### Supplemental Table 2f. Summary of HC participants by GFAP level.

|  | **GFAP tertiles** | | |  | | | | | |
| --- | --- | --- | --- | --- | --- | --- | --- | --- | --- |
| **Variable** | **<64.47757262** (N = 45) | **64.47757262-105.3411683** (N = 30) | **>105.3411683** (N = 22) | **Overall P-value** | **T1 vs. T2 P-value** | **T1 vs. T3 P-value** | **T2 vs. T3 P-value** | **Beta / OR (95% CI)** | **Continuous biomarker P-value** |
| **Sex**, n (%) |  |  |  | 0.2030 |  |  |  |  |  |
| Male | 26 (58%) | 21 (70%) | 10 (45%) |  |  |  |  |  |  |
| Female | 19 (42%) | 9 (30%) | 12 (55%) |  |  |  |  |  |  |
| **Age (years)**, mean (SD) | 52.9 (11.6) | 59.9 (10.3) | 69.8 (8.4) | **<0.0001** | **0.0087** | **<0.0001** | **0.0004** |  |  |
| Median (min, max) | 54.7 (30.6, 76.6) | 60.7 (31.9, 79.2) | 70.0 (50.3, 81.3) |  |  |  |  |  |  |
| Missing | 45 | 30 | 22 |  |  |  |  |  |  |
| **Race**, n (%) |  |  |  | 0.6701 |  |  |  |  |  |
| White | 41 (91%) | 26 (87%) | 21 (95%) |  |  |  |  |  |  |
| Black/African American | 4 (9%) | 2 (7%) | 0 |  |  |  |  |  |  |
| Asian | 0 | 0 | 1 (5%) |  |  |  |  |  |  |
| Other | 0 | 2 (7%) | 0 |  |  |  |  |  |  |
| **Ethnicity**, n (%) |  |  |  | 0.1430 |  |  |  |  |  |
| Not Hispanic/Latino | 45 (100%) | 28 (93%) | 22 (100%) |  |  |  |  |  |  |
| Hispanic/Latino | 0 | 2 (7%) | 0 |  |  |  |  |  |  |
| **UPSIT Raw Score**, mean (SD) | 35.0 (3.1) | 33.9 (4.0) | 33.4 (4.3) | 0.8249 ^f^ |  |  |  | 0.1071(-0.0052,0.2194) | 0.0614 ^f^ |
| Median (min, max) | 36.0 (27.0, 39.0) | 34.5 (21.0, 39.0) | 33.5 (21.0, 40.0) |  |  |  |  |  |  |
| **Revised UPSIT percentile**, mean (SD) | 60.8 (28.6) | 58.7 (29.2) | 59.9 (28.7) | 0.9799 |  |  |  | 0.0570(-0.0417,0.1557) | 0.2542 |
| Median (min, max) | 66.5 (2.0, 97.5) | 64.0 (3.0, 97.5) | 55.3 (6.0, 100.0) |  |  |  |  |  |  |
| **Revised UPSIT percentile ≤ 15**, n (%) | 5 (11%) | 4 (13%) | 1 (5%) | 0.6297 |  |  |  | 1.0066(0.9903,1.0232) | 0.4283 |
| **MDS-UPDRS item 1.1**, n (%) |  |  |  | 0.7051 ^a^ |  |  |  | 1.0234(0.9860,1.0622) | 0.2228 ^a^ |
| Normal | 40 (91%) | 30 (100%) | 21 (95%) |  |  |  |  |  |  |
| Slight | 3 (7%) | 0 | 1 (5%) |  |  |  |  |  |  |
| Mild | 1 (2%) | 0 | 0 |  |  |  |  |  |  |
| Missing | 1 | 0 | 0 |  |  |  |  |  |  |
| **MDS-UPDRS part I**, mean (SD) | 2.8 (2.9) | 1.7 (2.3) | 3.8 (2.9) | 0.6515 ^c^ |  |  |  | 0.0007(-0.0024,0.0038) | 0.6681 ^c^ |
| Median (min, max) | 2 (0, 12) | 1 (0, 8) | 3 (0, 11) |  |  |  |  |  |  |
| Missing | 1 | 0 | 0 |  |  |  |  |  |  |
| **MDS-UPDRS part II**, mean (SD) | 0.5 (1.1) | 0.6 (1.3) | 0.7 (0.8) | 0.1814 ^c^ |  |  |  | -0.0014(-0.0058,0.0030) | 0.5259 ^c^ |
| Median (min, max) | 0 (0, 6) | 0 (0, 5) | 0 (0, 2) |  |  |  |  |  |  |
| Missing | 1 | 0 | 0 |  |  |  |  |  |  |
| **MDS-UPDRS part III**, mean (SD) | 0.5 (0.8) | 1.3 (2.3) | 1.5 (3.3) | 0.3342 ^c^ |  |  |  | 0.0036(-0.0009,0.0081) | 0.1150 ^c^ |
| Median (min, max) | 0 (0, 3) | 0 (0, 8) | 0 (0, 13) |  |  |  |  |  |  |
| Missing | 1 | 0 | 0 |  |  |  |  |  |  |
| **MDS-UPDRS total score**, mean (SD) | 3.8 (3.9) | 3.6 (4.3) | 6.0 (4.6) | 0.7305 ^c^ |  |  |  | 0.0012(-0.0022,0.0046) | 0.4848 ^c^ |
| Median (min, max) | 3 (0, 15) | 2 (0, 16) | 5 (0, 17) |  |  |  |  |  |  |
| Missing | 1 | 0 | 0 |  |  |  |  |  |  |
| **MoCA total score**, mean (SD) | 28.5 (1.2) | 28.3 (1.1) | 28.2 (1.2) | 0.9831 ^f^ |  |  |  | -0.0109(-0.1255,0.1037) | 0.8510 ^f^ |
| Median (min, max) | 29 (27, 30) | 28 (27, 30) | 28 (27, 30) |  |  |  |  |  |  |
| **SCOPA-AUT total score**, mean (SD) | 5.0 (3.5) | 4.9 (3.0) | 8.2 (4.0) | 0.1726 ^c^ |  |  |  | 0.0006(-0.0024,0.0036) | 0.6937 ^d^ |
| Median (min, max) | 4 (0, 18) | 5 (1, 11) | 8 (2, 17) |  |  |  |  |  |  |
| **% Age/sex-expected low putamen**, mean (SD) | 0.99 (0.14) | 1.00 (0.14) | 1.01 (0.18) | 0.9000 |  |  |  | 0.0004(-0.0001,0.0009) | 0.1030 |
| Median (min, max) | 0.98 (0.65, 1.30) | 1.00 (0.71, 1.24) | 1.02 (0.68, 1.35) |  |  |  |  |  |  |
| Missing | 0 | 2 | 0 |  |  |  |  |  |  |
| **Mean striatum CWM**, mean (SD) | 1.49 (0.25) | 1.39 (0.22) | 1.36 (0.27) | 0.9148 ^b^ |  |  |  | 0.0007(-0.0002,0.0015) | 0.1330 ^b^ |
| Median (min, max) | 1.46 (1.02, 2.17) | 1.35 (1.08, 2.10) | 1.33 (0.96, 2.01) |  |  |  |  |  |  |
| Missing | 0 | 2 | 0 |  |  |  |  |  |  |
| **Mean putamen CWM**, mean (SD) | 1.58 (0.25) | 1.54 (0.22) | 1.57 (0.30) | 0.8694 ^b^ |  |  |  | 0.0010(<0.0001,0.0019) | **0.0496 ^b^** |
| Median (min, max) | 1.52 (1.00, 2.22) | 1.53 (1.17, 2.13) | 1.57 (0.99, 2.15) |  |  |  |  |  |  |
| Missing | 0 | 2 | 0 |  |  |  |  |  |  |
| **Cognitive categorization**, n (%) |  |  |  |  |  |  |  |  |  |
| Normal cognition | 5 (100%) | 4 (100%) | 6 (100%) |  |  |  |  |  |  |
| Missing | 40 | 26 | 16 |  |  |  |  |  |  |

#### Supplemental Table 2g. Summary of sPD participants by Aβ40 level.

|  | **Aβ40 plasma tertiles** | | |  | | | | | |
| --- | --- | --- | --- | --- | --- | --- | --- | --- | --- |
| **Variable** | **<374.886** (N = 37) | **374.886-422.785** (N = 40) | **>422.785** (N = 23) | **Overall P-value** | **T1 vs. T2 P-value** | **T1 vs. T3 P-value** | **T2 vs. T3 P-value** | **Beta / OR (95% CI)** | **Continuous biomarker P-value** |
| **Sex**, n (%) |  |  |  | 0.5823 |  |  |  |  |  |
| Male | 19 (51%) | 25 (63%) | 14 (61%) |  |  |  |  |  |  |
| Female | 18 (49%) | 15 (38%) | 9 (39%) |  |  |  |  |  |  |
| **Age (years)**, mean (SD) | 54.8 (8.8) | 61.1 (8.5) | 65.4 (8.2) | **<0.0001** | **0.0020** | **<0.0001** | 0.0732 |  |  |
| Median (min, max) | 55.3 (33.7, 78.0) | 61.2 (39.2, 77.3) | 65.2 (50.7, 84.9) |  |  |  |  |  |  |
| **Duration (years)**, mean (SD) | 0.6 (0.5) | 0.5 (0.4) | 0.5 (0.6) | 0.3373 |  |  |  |  |  |
| Median (min, max) | 0.4 (0.1, 2.6) | 0.3 (0.1, 1.8) | 0.3 (0.1, 2.7) |  |  |  |  |  |  |
| **Race**, n (%) |  |  |  | 0.0589 |  |  |  |  |  |
| White | 32 (89%) | 40 (100%) | 22 (96%) |  |  |  |  |  |  |
| Black/African American | 0 | 0 | 1 (4%) |  |  |  |  |  |  |
| Asian | 3 (8%) | 0 | 0 |  |  |  |  |  |  |
| Other | 1 (3%) | 0 | 0 |  |  |  |  |  |  |
| Missing | 1 | 0 | 0 |  |  |  |  |  |  |
| **Ethnicity**, n (%) |  |  |  | 1.0000 |  |  |  |  |  |
| Not Hispanic/Latino | 36 (97%) | 38 (95%) | 22 (96%) |  |  |  |  |  |  |
| Hispanic/Latino | 1 (3%) | 2 (5%) | 1 (4%) |  |  |  |  |  |  |
| **UPSIT Raw Score**, mean (SD) | 24.4 (6.9) | 23.3 (7.3) | 20.5 (7.2) | 0.5190 ^b^ |  |  |  | -0.0164(-0.0421,0.0092) | 0.2063 ^b^ |
| Median (min, max) | 24.0 (7.0, 36.0) | 23.5 (11.0, 38.0) | 20.0 (8.0, 36.0) |  |  |  |  |  |  |
| **Revised UPSIT percentile**, mean (SD) | 9.6 (12.1) | 13.2 (17.2) | 12.3 (15.7) | 0.4049 |  |  |  | 0.0099(-0.0412,0.0609) | 0.7023 |
| Median (min, max) | 6.0 (1.0, 60.5) | 8.5 (1.0, 91.5) | 6.0 (1.0, 60.5) |  |  |  |  |  |  |
| **Revised UPSIT percentile ≤ 15**, n (%) | 30 (81%) | 30 (75%) | 19 (83%) | 0.7179 |  |  |  | 1.0001(0.9920,1.0084) | 0.9716 |
| **NSD+**, n (%) |  |  |  | 0.0701 ^a^ |  |  |  | 0.9894(0.9799,0.9990) | **0.0306 ^a^** |
| 2a | 0 | 1 (3%) | 0 |  |  |  |  |  |  |
| 2b | 10 (27%) | 8 (20%) | 10 (43%) |  |  |  |  |  |  |
| 3 | 26 (70%) | 28 (70%) | 11 (48%) |  |  |  |  |  |  |
| 4 | 1 (3%) | 3 (8%) | 2 (9%) |  |  |  |  |  |  |
| **MDS-UPDRS item 1.1**, n (%) |  |  |  | 0.3899 ^a^ |  |  |  | 1.0016(0.9919,1.0114) | 0.7451 ^a^ |
| Normal | 31 (84%) | 32 (80%) | 21 (91%) |  |  |  |  |  |  |
| Slight | 6 (16%) | 5 (13%) | 2 (9%) |  |  |  |  |  |  |
| Mild | 0 | 3 (8%) | 0 |  |  |  |  |  |  |
| **MDS-UPDRS part I**, mean (SD) | 4.7 (3.2) | 5.5 (4.3) | 3.4 (2.6) | **0.0204 ^c^** | 0.8741 ^c^ | **0.0198 ^c^** | **0.0072 ^c^** | -0.0032(-0.0061,-0.0004) | **0.0260 ^c^** |
| Median (min, max) | 4 (0, 14) | 5 (0, 17) | 2 (0, 8) |  |  |  |  |  |  |
| **MDS-UPDRS part II**, mean (SD) | 5.1 (3.3) | 5.3 (3.4) | 4.5 (4.4) | 0.0522 ^c^ |  |  |  | -0.0029(-0.0056,-0.0002) | **0.0357 ^c^** |
| Median (min, max) | 4 (1, 13) | 5 (1, 15) | 3 (1, 18) |  |  |  |  |  |  |
| **MDS-UPDRS part III**, mean (SD) | 18.8 (8.1) | 18.4 (8.3) | 20.0 (7.4) | 0.6229 ^d^ |  |  |  | -0.0008(-0.0042,0.0026) | 0.6269 ^d^ |
| Median (min, max) | 20 (6, 40) | 17 (4, 41) | 18 (12, 40) |  |  |  |  |  |  |
| **MDS-UPDRS total score**, mean (SD) | 28.6 (11.0) | 29.2 (11.4) | 27.8 (10.9) | 0.4773 ^b^ |  |  |  | -0.0217(-0.0622,0.0189) | 0.2908 ^b^ |
| Median (min, max) | 30 (9, 50) | 29 (7, 53) | 26 (14, 62) |  |  |  |  |  |  |
| **MoCA total score**, mean (SD) | 27.8 (2.3) | 27.3 (2.0) | 26.7 (2.9) | 0.7928 ^f^ |  |  |  | -0.0387(-0.1427,0.0653) | 0.4621 ^f^ |
| Median (min, max) | 28 (21, 30) | 28 (21, 30) | 27 (21, 30) |  |  |  |  |  |  |
| **SCOPA-AUT total score**, mean (SD) | 9.1 (5.7) | 8.6 (6.6) | 8.8 (4.8) | 0.1961 ^d^ |  |  |  | -0.0028(-0.0064,0.0008) | 0.1244 ^d^ |
| Median (min, max) | 8 (2, 28) | 7 (0, 27) | 8 (2, 21) |  |  |  |  |  |  |
| **% Age/sex-expected low putamen**, mean (SD) | 0.38 (0.10) | 0.42 (0.13) | 0.44 (0.15) | 0.2350 |  |  |  | 0.0004(-0.0000,0.0008) | 0.0789 |
| Median (min, max) | 0.38 (0.19, 0.70) | 0.40 (0.20, 0.80) | 0.44 (0.22, 0.75) |  |  |  |  |  |  |
| **Mean striatum CWM**, mean (SD) | 0.87 (0.21) | 0.85 (0.18) | 0.81 (0.18) | 0.9238 ^b^ |  |  |  | 0.0002(-0.0005,0.0008) | 0.6543 ^b^ |
| Median (min, max) | 0.86 (0.50, 1.32) | 0.84 (0.48, 1.25) | 0.80 (0.42, 1.12) |  |  |  |  |  |  |
| **Mean putamen CWM**, mean (SD) | 0.75 (0.18) | 0.77 (0.20) | 0.77 (0.21) | 0.8979 ^b^ |  |  |  | 0.0004(-0.0004,0.0011) | 0.3205 ^b^ |
| Median (min, max) | 0.71 (0.45, 1.15) | 0.77 (0.44, 1.27) | 0.80 (0.43, 1.12) |  |  |  |  |  |  |
| **Cognitive categorization**, n (%) |  |  |  |  |  |  |  |  |  |
| Normal cognition | 13 (100%) | 12 (100%) | 7 (100%) | NA |  |  |  | NA | NA |
| Missing | 24 | 28 | 16 |  |  |  |  |  |  |

#### Supplemental Table 2h. Summary of sPD participants by Aβ42 level.

|  | **Aβ42 plasma tertiles** | | |  | | | | | |
| --- | --- | --- | --- | --- | --- | --- | --- | --- | --- |
| **Variable** | **<40.219** (N = 32) | **40.219-45.87** (N = 37) | **>45.87** (N = 31) | **Overall P-value** | **T1 vs. T2 P-value** | **T1 vs. T3 P-value** | **T2 vs. T3 P-value** | **Beta / OR (95% CI)** | **Continuous biomarker P-value** |
| **Sex**, n (%) |  |  |  | 0.4263 |  |  |  |  |  |
| Male | 20 (63%) | 23 (62%) | 15 (48%) |  |  |  |  |  |  |
| Female | 12 (38%) | 14 (38%) | 16 (52%) |  |  |  |  |  |  |
| **Age (years)**, mean (SD) | 55.6 (9.1) | 61.4 (7.9) | 62.0 (10.3) | **0.0162** | **0.0121** | **0.0147** | 0.7118 |  |  |
| Median (min, max) | 57.1 (33.7, 78.0) | 61.6 (47.5, 77.3) | 62.2 (39.2, 84.9) |  |  |  |  |  |  |
| **Duration (years)**, mean (SD) | 0.6 (0.5) | 0.5 (0.5) | 0.5 (0.5) | 0.4657 |  |  |  |  |  |
| Median (min, max) | 0.4 (0.1, 2.6) | 0.3 (0.1, 1.8) | 0.3 (0.1, 2.7) |  |  |  |  |  |  |
| **Race**, n (%) |  |  |  | 0.5153 |  |  |  |  |  |
| White | 28 (90%) | 36 (97%) | 30 (97%) |  |  |  |  |  |  |
| Black/African American | 0 | 0 | 1 (3%) |  |  |  |  |  |  |
| Asian | 2 (6%) | 1 (3%) | 0 |  |  |  |  |  |  |
| Other | 1 (3%) | 0 | 0 |  |  |  |  |  |  |
| Missing | 1 | 0 | 0 |  |  |  |  |  |  |
| **Ethnicity**, n (%) |  |  |  | 0.8315 |  |  |  |  |  |
| Not Hispanic/Latino | 30 (94%) | 36 (97%) | 30 (97%) |  |  |  |  |  |  |
| Hispanic/Latino | 2 (6%) | 1 (3%) | 1 (3%) |  |  |  |  |  |  |
| **UPSIT Raw Score**, mean (SD) | 24.7 (7.9) | 22.9 (6.9) | 21.5 (6.8) | 0.3974 ^b^ |  |  |  | -0.0793(-0.2697,0.1111) | 0.4105^b^ |
| Median (min, max) | 27.5 (7.0, 38.0) | 24.0 (11.0, 36.0) | 22.0 (8.0, 36.0) |  |  |  |  |  |  |
| **Revised UPSIT percentile**, mean (SD) | 13.2 (18.7) | 11.5 (12.3) | 10.2 (14.2) | 0.4703 |  |  |  | -0.1616(-0.5607,0.2375) | 0.4237 |
| Median (min, max) | 7.3 (1.0, 91.5) | 9.0 (1.0, 62.5) | 5.0 (1.0, 60.5) |  |  |  |  |  |  |
| **Revised UPSIT percentile ≤ 15**, n (%) | 26 (81%) | 28 (76%) | 25 (81%) | 0.8209 |  |  |  | 1.0168(0.9551,1.0825) | 0.6017 |
| **NSD+**, n (%) |  |  |  | 0.7599 ^a^ |  |  |  | 0.9924(0.9358,1.0524) | 0.7989 ^a^ |
| 2a | 1 (3%) | 0 | 0 |  |  |  |  |  |  |
| 2b | 9 (28%) | 9 (24%) | 10 (32%) |  |  |  |  |  |  |
| 3 | 21 (66%) | 26 (70%) | 18 (58%) |  |  |  |  |  |  |
| 4 | 1 (3%) | 2 (5%) | 3 (10%) |  |  |  |  |  |  |
| **MDS-UPDRS item 1.1**, n (%) |  |  |  | 0.3790 ^a^ |  |  |  | 1.0540(0.9681,1.1476) | 0.2254 ^a^ |
| Normal | 26 (81%) | 30 (81%) | 28 (90%) |  |  |  |  |  |  |
| Slight | 6 (19%) | 4 (11%) | 3 (10%) |  |  |  |  |  |  |
| Mild | 0 | 3 (8%) | 0 |  |  |  |  |  |  |
| **MDS-UPDRS part I**, mean (SD) | 4.3 (3.1) | 5.2 (4.0) | 4.4 (3.6) | 0.0835 ^c^ |  |  |  | -0.0092(-0.0304,0.0120) | 0.3916 ^c^ |
| Median (min, max) | 4 (0, 14) | 4 (0, 13) | 4 (0, 17) |  |  |  |  |  |  |
| **MDS-UPDRS part II**, mean (SD) | 5.0 (3.0) | 5.0 (3.4) | 5.3 (4.5) | 0.7845 ^c^ |  |  |  | 0.0198(-0.7772,0.8168) | 0.9608 ^f^ |
| Median (min, max) | 5 (1, 13) | 5 (1, 14) | 4 (1, 18) |  |  |  |  |  |  |
| **MDS-UPDRS part III**, mean (SD) | 17.0 (7.1) | 19.9 (7.9) | 19.7 (8.7) | 0.5742 ^d^ |  |  |  | 0.1084(-0.1062,0.3229) | 0.3185 ^b^ |
| Median (min, max) | 18 (4, 28) | 18 (5, 41) | 18 (7, 40) |  |  |  |  |  |  |
| **MDS-UPDRS total score**, mean (SD) | 26.3 (9.6) | 30.1 (10.6) | 29.3 (12.7) | 0.6354 ^b^ |  |  |  | 0.2110(-0.0873,0.5094) | 0.1636 ^b^ |
| Median (min, max) | 28 (9, 43) | 31 (7, 53) | 26 (11, 62) |  |  |  |  |  |  |
| **MoCA total score**, mean (SD) | 28.0 (2.1) | 26.8 (2.3) | 27.3 (2.5) | 0.1730 ^f^ |  |  |  | -0.2160(-0.9859,0.5539) | 0.5789 ^f^ |
| Median (min, max) | 29 (21, 30) | 27 (21, 30) | 28 (21, 30) |  |  |  |  |  |  |
| **SCOPA-AUT total score**, mean (SD) | 9.1 (5.9) | 8.9 (6.7) | 8.5 (4.7) | 0.3329 ^d^ |  |  |  | -0.0056(-0.0326,0.0214) | 0.6808 ^d^ |
| Median (min, max) | 8 (2, 28) | 7 (0, 27) | 8 (0, 21) |  |  |  |  |  |  |
| **% Age/sex-expected low putamen**, mean (SD) | 0.41 (0.13) | 0.42 (0.12) | 0.41 (0.13) | 0.9256 |  |  |  | -0.0008(-0.0041,0.0026) | 0.6560 |
| Median (min, max) | 0.41 (0.19, 0.80) | 0.40 (0.20, 0.75) | 0.38 (0.20, 0.68) |  |  |  |  |  |  |
| **Mean striatum CWM**, mean (SD) | 0.88 (0.21) | 0.84 (0.17) | 0.82 (0.19) | 0.7010 ^b^ |  |  |  | -0.0028(-0.0079,0.0022) | 0.2695 ^b^ |
| Median (min, max) | 0.91 (0.50, 1.32) | 0.83 (0.48, 1.25) | 0.78 (0.42, 1.25) |  |  |  |  |  |  |
| **Mean putamen CWM**, mean (SD) | 0.76 (0.21) | 0.77 (0.18) | 0.76 (0.20) | 0.9543 ^b^ |  |  |  | -0.0015(-0.0068,0.0039) | 0.5942 ^b^ |
| Median (min, max) | 0.74 (0.45, 1.27) | 0.76 (0.45, 1.15) | 0.72 (0.43, 1.11) |  |  |  |  |  |  |
| **Cognitive categorization**, n (%) |  |  |  | NA |  |  |  | NA | NA |
| Normal cognition | 14 (100%) | 10 (100%) | 8 (100%) |  |  |  |  |  |  |
| Missing | 18 | 27 | 23 |  |  |  |  |  |  |

#### Supplemental Table 2i. Summary of sPD participants by Aβ42/Aβ40 level

|  | **Aβ42/Aβ40 plasma tertiles** | | |  | | | | | |
| --- | --- | --- | --- | --- | --- | --- | --- | --- | --- |
| **Variable** | **<0.10402** (N = 26) | **0.10402-0.110585** (N = 29) | **>0.110585** (N = 45) | **Overall P-value** | **T1 vs. T2 P-value** | **T1 vs. T3 P-value** | **T2 vs. T3 P-value** | **Beta / OR (95% CI)** | **Continuous biomarker P-value** |
| **Sex**, n (%) |  |  |  | **0.0036** | 0.5083 | **0.0027** | **0.0149** |  |  |
| Male | 20 (77%) | 20 (69%) | 18 (40%) |  |  |  |  |  |  |
| Female | 6 (23%) | 9 (31%) | 27 (60%) |  |  |  |  |  |  |
| **Age (years)**, mean (SD) | 64.3 (8.6) | 58.9 (9.1) | 57.6 (9.4) | **0.0226** | **0.0244** | **0.0108** | 0.6943 |  |  |
| Median (min, max) | 64.5 (48.4, 84.9) | 59.1 (39.2, 78.0) | 57.4 (33.7, 73.9) |  |  |  |  |  |  |
| **Duration (years)**, mean (SD) | 0.5 (0.3) | 0.5 (0.5) | 0.6 (0.6) | 0.5808 |  |  |  |  |  |
| Median (min, max) | 0.4 (0.1, 1.5) | 0.4 (0.1, 1.8) | 0.3 (0.1, 2.7) |  |  |  |  |  |  |
| **Race**, n (%) |  |  |  | 0.5149 |  |  |  |  |  |
| White | 26 (100%) | 27 (93%) | 41 (93%) |  |  |  |  |  |  |
| Black/African American | 0 | 0 | 1 (2%) |  |  |  |  |  |  |
| Asian | 0 | 2 (7%) | 1 (2%) |  |  |  |  |  |  |
| Other | 0 | 0 | 1 (2%) |  |  |  |  |  |  |
| Missing | 0 | 0 | 1 |  |  |  |  |  |  |
| **Ethnicity**, n (%) |  |  |  | 0.6885 |  |  |  |  |  |
| Not Hispanic/Latino | 24 (92%) | 28 (97%) | 44 (98%) |  |  |  |  |  |  |
| Hispanic/Latino | 2 (8%) | 1 (3%) | 1 (2%) |  |  |  |  |  |  |
| **UPSIT Raw Score**, mean (SD) | 21.0 (8.3) | 25.0 (6.9) | 22.9 (6.6) | 0.1500 ^b^ |  |  |  | 15.8587(-63.8540,95.5714) | 0.6938 ^b^ |
| Median (min, max) | 19.5 (8.0, 38.0) | 26.0 (7.0, 36.0) | 24.0 (11.0, 36.0) |  |  |  |  |  |  |
| **Revised UPSIT percentile**, mean (SD) | 15.0 (22.0) | 13.6 (12.7) | 8.5 (10.7) | **0.0407** | 0.1739 | 0.3151 | **0.0125** | -93.1103(-257.060,70.8400) | 0.2625 |
| Median (min, max) | 5.0 (1.0, 91.5) | 12.0 (1.0, 62.5) | 5.0 (1.0, 60.5) |  |  |  |  |  |  |
| **Revised UPSIT percentile ≤ 15**, n (%) | 21 (81%) | 20 (69%) | 38 (84%) | 0.2707 |  |  |  | 19538.59(0.0000,3.332E14) | 0.4111 |
| **NSD+**, n (%) |  |  |  | **0.0198 ^a^** | **0.0296 ^a^** | **0.0074 ^a^** | 0.6073 ^a^ | >99.9999(0.0003,>99.9999) | 0.1119 ^a^ |
| 2a | 1 (4%) | 0 | 0 |  |  |  |  |  |  |
| 2b | 11 (42%) | 7 (24%) | 10 (22%) |  |  |  |  |  |  |
| 3 | 14 (54%) | 19 (66%) | 32 (71%) |  |  |  |  |  |  |
| 4 | 0 | 3 (10%) | 3 (7%) |  |  |  |  |  |  |
| **MDS-UPDRS item 1.1**, n (%) |  |  |  | 0.9271 ^a^ |  |  |  | >99.9999(<0.0001,>99.9999) | 0.2307 ^a^ |
| Normal | 21 (81%) | 25 (86%) | 38 (84%) |  |  |  |  |  |  |
| Slight | 5 (19%) | 3 (10%) | 5 (11%) |  |  |  |  |  |  |
| Mild | 0 | 1 (3%) | 2 (4%) |  |  |  |  |  |  |
| **MDS-UPDRS part I**, mean (SD) | 3.8 (2.9) | 5.1 (3.9) | 5.0 (3.7) | 0.3809 ^d^ |  |  |  | 8.8090(-2.0263,19.6442) | 0.1099 ^d^ |
| Median (min, max) | 4 (0, 10) | 4 (0, 14) | 4 (0, 17) |  |  |  |  |  |  |
| **MDS-UPDRS part II**, mean (SD) | 3.9 (2.8) | 5.1 (3.6) | 5.7 (4.0) | **0.0215 ^c^** | **0.0426 ^c^** | **0.0065 ^c^** | 0.4799 ^c^ | 9.0922(0.8250,17.3593) | **0.0315 ^c^** |
| Median (min, max) | 4 (1, 11) | 4 (1, 14) | 5 (1, 18) |  |  |  |  |  |  |
| **MDS-UPDRS part III**, mean (SD) | 17.7 (6.6) | 19.0 (7.9) | 19.6 (8.8) | 0.2188 ^d^ |  |  |  | 84.7434(-3.6560,173.1428) | 0.0601 ^b^ |
| Median (min, max) | 17 (4, 31) | 18 (5, 41) | 18 (7, 41) |  |  |  |  |  |  |
| **MDS-UPDRS total score**, mean (SD) | 25.4 (9.8) | 29.2 (10.3) | 30.2 (12.0) | **0.0384 ^b^** | 0.0612 ^b^ | **0.0123 ^b^** | 0.5224 ^b^ | 170.2536(49.2036,291.3035) | **0.0063 ^b^** |
| Median (min, max) | 24 (10, 41) | 31 (7, 53) | 29 (9, 62) |  |  |  |  |  |  |
| **MoCA total score**, mean (SD) | 26.5 (2.8) | 28.0 (1.7) | 27.4 (2.3) | 0.3204 ^f^ |  |  |  | 17.4697(-304.4957,339.4351) | 0.9145 ^f^ |
| Median (min, max) | 27 (21, 30) | 28 (25, 30) | 28 (21, 30) |  |  |  |  |  |  |
| **SCOPA-AUT total score**, mean (SD) | 8.8 (5.6) | 9.4 (6.3) | 8.5 (5.8) | 0.6715 ^d^ |  |  |  | 4.9330(-6.2986,16.1647) | 0.3855 ^d^ |
| Median (min, max) | 8 (2, 21) | 8 (0, 28) | 7 (0, 27) |  |  |  |  |  |  |
| **% Age/sex-expected low putamen**, mean (SD) | 0.44 (0.14) | 0.43 (0.14) | 0.38 (0.10) | 0.0727 |  |  |  | -1.5270(-2.8672,-0.1869) | **0.0260** |
| Median (min, max) | 0.45 (0.22, 0.80) | 0.43 (0.20, 0.75) | 0.36 (0.19, 0.61) |  |  |  |  |  |  |
| **Mean striatum CWM**, mean (SD) | 0.83 (0.21) | 0.89 (0.19) | 0.82 (0.18) | 0.0827 ^b^ |  |  |  | -2.0171(-4.1133,0.0792) | 0.0591 ^b^ |
| Median (min, max) | 0.81 (0.42, 1.25) | 0.90 (0.54, 1.32) | 0.78 (0.48, 1.25) |  |  |  |  |  |  |
| **Mean putamen CWM**, mean (SD) | 0.80 (0.22) | 0.79 (0.20) | 0.73 (0.17) | 0.1623 ^b^ |  |  |  | -1.8685(-4.0896,0.3525) | 0.0982 ^b^ |
| Median (min, max) | 0.79 (0.43, 1.27) | 0.81 (0.45, 1.15) | 0.71 (0.43, 1.06) |  |  |  |  |  |  |
| **Cognitive categorization**, n (%) |  |  |  | NA |  |  |  | NA | NA |
| Normal cognition | 4 (100%) | 13 (100%) | 15 (100%) |  |  |  |  |  |  |
| Missing | 22 | 16 | 30 |  |  |  |  |  |  |

Notes: Report Generated on Data Submitted as of: 08DEC2025.

^a^ P-values calculated using PROC LOGISTIC adjusted for age and sex.
^b^ P-values calculated using GLM adjusted for age and sex.
^c^ P-values calculated using a log-transformed model.
^d^ P-values calculated using a square-root-transformed model.
^e^ P-values calculated using a squared-value (power-2) transformation.
^f^ P-values calculated using a rank-based nonparametric model.

No superscript indicate unadjusted p-values. Kruskal-Wallis and Chi-Square/Fisher's Exact Test are used for overall p-values. Wilcoxon Rank Sum tests and Chi-Square/Fisher's Exact Test were used for pairwise comparisons.

In p-value calculations, the race variable collapsed all non-white groups together and the resulting p-value is a comparison of non-white vs. white between groups.

For MDS-UPDRS item 1.1 and COGSTATE, all groups except 'Normal' are collapsed and the resulting p-value compares normal vs. abnormal categories.

For NSD Stage, stages 1-2.5 are collapsed and compared against stages 3-6, where applicable.

Beta / OR (95% CI) and continuous biomarker p-values are from models using the table biomarker as a continuous predictor. Unless otherwise specified, models are adjusted for age and sex. UPSIT percentile, UPSIT percentile <=15, and % age/sex-expected low putamen models are unadjusted.

#### Supplemental Table 3a: All Continuous sPD Mixed Models Summary

| **Biomarker** | **Outcome** | **Coefficient (95% CI)** | **SE** | **T-stat** | **p-value** |
| --- | --- | --- | --- | --- | --- |
| **pTau217 plasma** | MDS-UPDRS I Score | 1.8079 (0.3195,3.2964) | 0.7589 | 2.38 | **0.0173** |
|  | MDS-UPDRS II Score | 2.6437 (0.5286,4.7588) | 1.0784 | 2.45 | **0.0143** |
|  | MDS-UPDRS III Score OFF | 1.9252 (-1.2748,5.1251) | 1.6310 | 1.18 | 0.2381 |
|  | MOCA | -1.7869 (-2.8065,-0.7673) | 0.5199 | -3.44 | **0.0006** |
|  | Lowest Putamen Ratio | -0.0148 (-0.0486,0.0190) | 0.0172 | -0.86 | 0.3881 |
| **NFL** | MDS-UPDRS I Score | 0.0223 (0.0030,0.0415) | 0.0098 | 2.27 | **0.0233** |
|  | MDS-UPDRS II Score | 0.0281 (0.0046,0.0517) | 0.0120 | 2.35 | **0.0191** |
|  | MDS-UPDRS III Score OFF | 0.0194 (-0.0208,0.0596) | 0.0205 | 0.95 | 0.3430 |
|  | MOCA | -0.0152 (-0.0284,-0.0021) | 0.0067 | -2.27 | **0.0235** |
|  | Lowest Putamen Ratio | -0.0002 (-0.0007,0.0004) | 0.0003 | -0.62 | 0.5340 |
| **GFAP** | MDS-UPDRS I Score | 0.0019 (-0.0004,0.0043) | 0.0012 | 1.60 | 0.1095 |
|  | MDS-UPDRS II Score | 0.0025 (-7.02999E-05,0.0052) | 0.0013 | 1.91 | 0.0565 |
|  | MDS-UPDRS III Score OFF | 0.0001 (-0.0046,0.0049) | 0.0024 | 0.06 | 0.9530 |
|  | MOCA | -0.0021 (-0.0035,-0.0007) | 0.0007 | -3.00 | **0.0027** |
|  | Lowest Putamen Ratio | 8.49380E-06 (-4.46104E-05,6.15980E-05) | 0.0000 | 0.32 | 0.7530 |
| **Aß40 plasma** | MDS-UPDRS I Score | -1.60663E-05 (-0.0020,0.0019) | 0.0010 | -0.02 | 0.9872 |
|  | MDS-UPDRS II Score | 0.0009 (-0.0009,0.0028) | 0.0010 | 1.00 | 0.3192 |
|  | MDS-UPDRS III Score OFF | 0.0002 (-0.0050,0.0054) | 0.0026 | 0.07 | 0.9422 |
|  | MOCA | -0.0002 (-0.0011,0.0007) | 0.0005 | -0.41 | 0.6848 |
|  | Lowest Putamen Ratio | -8.38973E-05 (-0.0002,-4.66278E-06) | 0.0000 | -2.10 | **0.0382** |
| **Aß42 plasma** | MDS-UPDRS I Score | -0.0091 (-0.0260,0.0077) | 0.0086 | -1.06 | 0.2878 |
|  | MDS-UPDRS II Score | -0.0018 (-0.0174,0.0138) | 0.0079 | -0.23 | 0.8179 |
|  | MDS-UPDRS III Score OFF | -0.0306 (-0.0650,0.0038) | 0.0175 | -1.75 | 0.0809 |
|  | MOCA | 2.62486E-05 (-0.0087,0.0087) | 0.0044 | 0.01 | 0.9953 |
|  | Lowest Putamen Ratio | -0.0004 (-0.0010,0.0003) | 0.0003 | -1.08 | 0.2845 |
| **Aß42/ Aß40 plasma** | MDS-UPDRS I Score | -4.9767 (-11.0069,1.0535) | 3.0720 | -1.62 | 0.1056 |
|  | MDS-UPDRS II Score | -4.6561 (-9.4608,0.1486) | 2.4477 | -1.90 | 0.0575 |
|  | MDS-UPDRS III Score OFF | -15.8612 (-29.7053,-2.0171) | 7.0473 | -2.25 | **0.0248** |
| **Biomarker** | **Outcome** | **Coefficient (95% CI)** | **SE** | **T-stat** | **p-value** |
|  | MOCA | 0.8130 (-2.1132,3.7392) | 1.4906 | 0.55 | 0.5856 |
|  | Lowest Putamen Ratio | 0.0655 (-0.1524,0.2834) | 0.1096 | 0.60 | 0.5518 |

All Mixed models were adjusted for age and sex except for Lowest Putamen ratio.

Estimates and p-values are based on time and continuous biomarker interaction.

#### Supplemental Table 3b: All Continuous HC Mixed Models Summary

| **Biomarker** | **Outcome** | **Coefficient (95% CI)** | **SE** | **T-stat** | **p-value** |
| --- | --- | --- | --- | --- | --- |
| **pTau217** | MDS-UPDRS I Score | -0.3629 (-0.9770,0.2513) | 0.3126 | -1.16 | 0.2462 |
|  | MDS-UPDRS II Score | 0.2777 (-0.3959,0.9514) | 0.3428 | 0.81 | 0.4183 |
|  | MDS-UPDRS III Score OFF | 0.8943 (-0.8445,2.6330) | 0.8848 | 1.01 | 0.3127 |
|  | MOCA | -0.0938 (-0.4353,0.2477) | 0.1738 | -0.54 | 0.5897 |
| **NFL** | MDS-UPDRS I Score | 0.0043 (-0.0036,0.0121) | 0.0040 | 1.07 | 0.2849 |
|  | MDS-UPDRS II Score | 0.0104 (0.0006,0.0202) | 0.0050 | 2.09 | 0.0376 |
|  | MDS-UPDRS III Score OFF | 0.0345 (-0.0112,0.0803) | 0.0233 | 1.48 | 0.1383 |
|  | MOCA | -0.0026 (-0.0077,0.0025) | 0.0026 | -1.00 | 0.3160 |
| **GFAP** | MDS-UPDRS I Score | -0.0007 (-0.0020,0.0006) | 0.0007 | -1.06 | 0.2910 |
|  | MDS-UPDRS II Score | 0.0002 (-0.0005,0.0010) | 0.0004 | 0.60 | 0.5502 |
|  | MDS-UPDRS III Score OFF | 0.0026 (-0.0025,0.0077) | 0.0026 | 1.01 | 0.3127 |
|  | MOCA | -0.0007 (-0.0011,-0.0003) | 0.0002 | -3.34 | **0.0009** |

All Mixed models were adjusted for age and sex. Lowest Putamen ratio models were omitted due to low longitudinal counts for HCs.

Estimates and p-values are based on time and continuous biomarker interaction.

#### Supplement Table 3c: All Categorical sPD Mixed Model Summary

| **Biomarker** | **Outcome** | **Comparison** | **Coefficient (95% CI)** | **SE** | **F / t-stat** | **p-value** |
| --- | --- | --- | --- | --- | --- | --- |
| **pTau217 plasma** | MDS-UPDRS I Score | Overall |  |  | 1.76 | 0.1728 |
|  | MDS-UPDRS II Score | Overall |  |  | 3.12 | **0.0442** |
|  |  | T1 vs. T2 | -0.0227 (-0.2222,0.1768) | 0.1017 | -0.22 | 0.8236 |
|  |  | T1 vs. T3 | -0.3214 (-0.5810,-0.0619) | 0.1323 | -2.43 | **0.0152** |
|  |  | T2 vs. T3 | -0.2987 (-0.5705,-0.0269) | 0.1386 | -2.16 | **0.0312** |
|  | MDS-UPDRS III Score OFF | Overall |  |  | 0.85 | 0.4266 |
|  | MOCA | Overall |  |  | 3.99 | **0.0186** |
|  |  | T1 vs. T2 | 0.0171 (-0.0865,0.1207) | 0.0528 | 0.32 | 0.7461 |
|  |  | T1 vs. T3 | 0.1828 (0.0547,0.3109) | 0.0653 | 2.80 | **0.0052** |
|  |  | T2 vs. T3 | 0.1657 (0.0226,0.3088) | 0.0730 | 2.27 | **0.0233** |
|  | Lowest Putamen Ratio | Overall |  |  | 1.07 | 0.3460 |
| **NFL** | MDS-UPDRS I Score | Overall |  |  | 3.80 | **0.0226** |
|  |  | T1 vs. T2 | 0.0122 (-0.1599,0.1844) | 0.0878 | 0.14 | 0.8891 |
|  |  | T1 vs. T3 | -0.2693 (-0.4856,-0.0530) | 0.1103 | -2.44 | **0.0147** |
|  |  | T2 vs. T3 | -0.2815 (-0.4943,-0.0688) | 0.1085 | -2.60 | **0.0095** |
|  | MDS-UPDRS II Score | Overall |  |  | 6.47 | **0.0016** |
|  |  | T1 vs. T2 | -0.1325 (-0.3280,0.0630) | 0.0997 | -1.33 | 0.1840 |
|  |  | T1 vs. T3 | -0.4755 (-0.7350,-0.2160) | 0.1323 | -3.59 | **0.0003** |
|  |  | T2 vs. T3 | -0.3430 (-0.6117,-0.0743) | 0.1370 | -2.50 | **0.0124** |
|  | MDS-UPDRS III Score OFF | Overall |  |  | 0.48 | 0.6197 |
|  | MOCA | Overall |  |  | 6.17 | **0.0021** |
|  |  | T1 vs. T2 | 0.1014 (0.0124,0.1903) | 0.0454 | 2.23 | **0.0256** |
|  |  | T1 vs. T3 | 0.2340 (0.0877,0.3803) | 0.0746 | 3.14 | **0.0017** |
|  |  | T2 vs. T3 | 0.1326 (-0.0207,0.2860) | 0.0782 | 1.70 | 0.0900 |
|  | Lowest Putamen Ratio | Overall |  |  | 0.81 | 0.4451 |
| **GFAP** | MDS-UPDRS I Score | Overall |  |  | 0.36 | 0.6970 |
|  | MDS-UPDRS II Score | Overall |  |  | 1.61 | 0.2005 |
|  | MDS-UPDRS III Score OFF | Overall |  |  | 0.43 | 0.6504 |
|  | MOCA | Overall |  |  | 3.37 | **0.0345** |
| **Biomarker** | **Outcome** | **Comparison** | **Coefficient (95% CI)** | **SE** | **F / t-stat** | **p-value** |
|  |  | T1 vs. T2 | 0.0767 (-0.0197,0.1730) | 0.0491 | 1.56 | 0.1189 |
|  |  | T1 vs. T3 | 0.1706 (0.0378,0.3034) | 0.0677 | 2.52 | **0.0118** |
|  |  | T2 vs. T3 | 0.0939 (-0.0363,0.2242) | 0.0664 | 1.41 | 0.1573 |
|  | Lowest Putamen Ratio | Overall |  |  | 0.35 | 0.7026 |
| **Aß40 plasma** | MDS-UPDRS I Score | Overall |  |  | 0.06 | 0.9435 |
|  | MDS-UPDRS II Score | Overall |  |  | 0.41 | 0.6664 |
|  | MDS-UPDRS III Score OFF | Overall |  |  | 0.23 | 0.7970 |
|  | MOCA | Overall |  |  | 0.23 | 0.7983 |
|  | Lowest Putamen Ratio | Overall |  |  | 0.73 | 0.4853 |
| **Aß42 plasma** | MDS-UPDRS I Score | Overall |  |  | 1.65 | 0.1936 |
|  | MDS-UPDRS II Score | Overall |  |  | 1.05 | 0.3510 |
|  | MDS-UPDRS III Score OFF | Overall |  |  | 0.81 | 0.4451 |
|  | MOCA | Overall |  |  | 0.11 | 0.8917 |
|  | Lowest Putamen Ratio | Overall |  |  | 1.94 | 0.1495 |
| **Aß42/Aß40 plasma** | MDS-UPDRS I Score | Overall |  |  | 1.14 | 0.3212 |
|  | MDS-UPDRS II Score | Overall |  |  | 1.08 | 0.3409 |
|  | MDS-UPDRS III Score OFF | Overall |  |  | 0.76 | 0.4670 |
|  | MOCA | Overall |  |  | 1.49 | 0.2270 |
|  | Lowest Putamen Ratio | Overall |  |  | 0.35 | 0.7024 |

All Mixed models were adjusted for age and sex except for Lowest Putamen ratio. T3 was the reference group.

F-statistics were calculated for overall p-values and t-statistics were used for pairwise p-value calculations.

Overall p-values correspond to the omnibus Type III test for the time and tertile interaction term.

Coefficient, standard error, and 95% confidence interval are not shown for the Overall row because the omnibus test is a multi-degree-of-freedom test and does not yield a single parameter estimate.

#### Supplement Table 3d: All Categorical HC Mixed Model Summary

| **Biomarker** | **Outcome** | **Comparison** | **F / t-stat** | **p-value** |
| --- | --- | --- | --- | --- |
| **pTau217** | MDS-UPDRS I Score | Overall | 0.78 | 0.4581 |
|  | MDS-UPDRS II Score | Overall | 2.23 | 0.1087 |
|  | MDS-UPDRS III Score OFF | Overall | 1.75 | 0.1745 |
|  | MOCA | Overall | 0.13 | 0.8758 |
| **NFL** | MDS-UPDRS I Score | Overall | 0.12 | 0.8857 |
|  | MDS-UPDRS II Score | Overall | 1.16 | 0.3131 |
|  | MDS-UPDRS III Score OFF | Overall | 1.25 | 0.2875 |
|  | MOCA | Overall | 1.63 | 0.1975 |
| **GFAP** | MDS-UPDRS I Score | Overall | 0.54 | 0.5819 |
|  | MDS-UPDRS II Score | Overall | 2.30 | 0.1012 |
|  | MDS-UPDRS III Score OFF | Overall | 1.07 | 0.3424 |
|  | MOCA | Overall | 1.49 | 0.2255 |

All Mixed models were adjusted for age and sex. Lowest Putamen ratio models were omitted due to low longitudinal counts for HCs. T3 was the reference group.

F-statistics were calculated for overall p-values and t-statistics were used for pairwise p-value calculations.

Overall p-values correspond to the omnibus Type III test for the time and tertile interaction term.

#### Supplement Table 4a: All Continuous sPD Progression Milestones

| **Biomarker** | **Milestone** | **Model** | **Beta (95% CI)** | **SE** | **Z** | **p-value** | **HR / Derived HR (95% CI)** |
| --- | --- | --- | --- | --- | --- | --- | --- |
| pTau217 plasma | Any progression milestone | CoxPH | 1.859 (0.529,3.188) | 0.678 | 2.740 | **0.006** | 6.414 (1.697,24.241) |
|  | Walking and balance domain | AFT | -1.347 (-2.049,-0.645) | 0.358 | -3.759 | **<0.001** | 20.060 (4.106,98.002) |
|  | Motor complications domain | CoxPH | 1.035 (-1.484,3.555) | 1.285 | 0.805 | 0.421 | 2.816 (0.227,34.974) |
|  | Cognitive domain | CoxPH | 3.173 (1.697,4.650) | 0.753 | 4.213 | **<0.001** | 23.887 (5.458,104.544) |
|  | Autonomic dysfunction domain | CoxPH | 0.360 (-1.695,2.415) | 1.048 | 0.343 | 0.731 | 1.433 (0.184,11.190) |
|  | Functional dependence domain | AFT | -1.345 (-2.113,-0.577) | 0.392 | -3.433 | **<0.001** | 14.126 (3.051,65.397) |
| NFL | Any progression milestone | AFT | 0.004 (-0.012,0.020) | 0.008 | 0.490 | 0.624 | 0.994 (0.969,1.019) |
|  | Walking and balance domain | AFT | -0.005 (-0.021,0.011) | 0.008 | -0.647 | 0.518 | 1.012 (0.977,1.048) |
|  | Motor complications domain | CoxPH | -0.015 (-0.057,0.028) | 0.022 | -0.685 | 0.493 | 0.985 (0.944,1.028) |
|  | Cognitive domain | CoxPH | 0.018 (-0.014,0.050) | 0.016 | 1.108 | 0.268 | 1.018 (0.986,1.051) |
|  | Autonomic dysfunction domain | CoxPH | 0.013 (-0.022,0.047) | 0.018 | 0.711 | 0.477 | 1.013 (0.978,1.048) |
|  | Functional dependence domain | CoxPH | -0.023 (-0.059,0.013) | 0.018 | -1.273 | 0.203 | 0.977 (0.942,1.013) |
| GFAP | Any progression milestone | AFT | -0.001 (-0.002,0.001) | 0.001 | -0.727 | 0.467 | 1.001 (0.998,1.004) |
|  | Walking and balance domain | AFT | -0.002 (-0.004,-0.000) | 0.001 | -2.199 | **0.028** | 1.004 (1.000,1.008) |
|  | Motor complications domain | CoxPH | -0.003 (-0.008,0.001) | 0.002 | -1.451 | 0.147 | 0.997 (0.992,1.001) |
|  | Cognitive domain | CoxPH | 0.004 (0.001,0.007) | 0.002 | 2.420 | **0.016** | 1.004 (1.001,1.007) |
|  | Autonomic dysfunction domain | CoxPH | 0.003 (-0.000,0.007) | 0.002 | 1.818 | 0.069 | 1.003 (1.000,1.007) |
|  | Functional dependence domain | AFT | -0.000 (-0.002,0.002) | 0.001 | -0.027 | 0.978 | 1.000 (0.996,1.004) |
| Aβ40 plasma | Any progression milestone | CoxPH | 0.000 (-0.003,0.004) | 0.002 | 0.098 | 0.922 | 1.000 (0.997,1.004) |
|  | Walking and balance domain | AFT | -0.000 (-0.002,0.002) | 0.001 | -0.352 | 0.725 | 1.001 (0.996,1.006) |
|  | Motor complications domain | CoxPH | 0.002 (-0.003,0.007) | 0.003 | 0.673 | 0.501 | 1.002 (0.997,1.007) |
|  | Cognitive domain | CoxPH | -0.001 (-0.007,0.004) | 0.003 | -0.379 | 0.704 | 0.999 (0.993,1.004) |
|  | Autonomic dysfunction domain | CoxPH | 0.001 (-0.005,0.007) | 0.003 | 0.292 | 0.770 | 1.001 (0.995,1.007) |
|  | Functional dependence domain | CoxPH | 0.001 (-0.003,0.006) | 0.002 | 0.471 | 0.637 | 1.001 (0.997,1.006) |
| Aβ42 plasma | Any progression milestone | CoxPH | -0.002 (-0.035,0.031) | 0.017 | -0.119 | 0.906 | 0.998 (0.965,1.032) |
|  | Walking and balance domain | AFT | -0.013 (-0.029,0.003) | 0.008 | -1.560 | 0.119 | 1.037 (0.991,1.084) |
|  | Motor complications domain | CoxPH | 0.007 (-0.030,0.044) | 0.019 | 0.359 | 0.720 | 1.007 (0.970,1.045) |
|  | Cognitive domain | CoxPH | 0.019 (-0.033,0.071) | 0.026 | 0.726 | 0.468 | 1.019 (0.968,1.073) |
|  | Autonomic dysfunction domain | CoxPH | 0.027 (-0.019,0.074) | 0.024 | 1.156 | 0.248 | 1.028 (0.981,1.077) |
|  | Functional dependence domain | CoxPH | 0.004 (-0.037,0.045) | 0.021 | 0.187 | 0.852 | 1.004 (0.964,1.046) |
| Aβ42/Aβ40 plasma | Any progression milestone | CoxPH | -2.953 (-14.646,8.740) | 5.966 | -0.495 | 0.621 | 0.052 (0.000,6248.279) |
|  | Walking and balance domain | AFT | -3.368 (-8.651,1.915) | 2.695 | -1.249 | 0.212 | 10495.70 (0.006,1.804E10) |
|  | Motor complications domain | CoxPH | -4.602 (-21.466,12.262) | 8.604 | -0.535 | 0.593 | 0.010 (0.000,211438.3) |
|  | Cognitive domain | CoxPH | 8.458 (-8.144,25.060) | 8.470 | 0.999 | 0.318 | 4711.749 (0.000,7.643E10) |
|  | Autonomic dysfunction domain | CoxPH | 4.390 (-11.834,20.614) | 8.277 | 0.530 | 0.596 | 80.640 (0.000,8.9622E8) |
|  | Functional dependence domain | CoxPH | -6.673 (-24.677,11.332) | 9.186 | -0.726 | 0.468 | 0.001 (0.000,83420.58) |

***Cox PH models report HR per 1-unit increase in biomarkers. Weibull AFT models report derived HR using HR=exp(-beta/sigma). Models adjusted for age and sex.***

#### Supplement Table 4b: All Continuous sPD Clinical Milestones

| **Biomarker** | **Milestone** | **Model** | **Coefficient (95% CI)** | **SE** | **Z** | **p** | **HR (95% CI)** |
| --- | --- | --- | --- | --- | --- | --- | --- |
| pTau217 plasma | Time to HY >= 3 | CoxPH | **2.247 (0.731,3.764)** | 0.774 | 2.904 | **0.004** | **9.461 (2.076,43.111)** |
|  | Time to Cognitive State Change | CoxPH | **5.135 (1.717,8.553)** | 1.744 | 2.944 | **0.003** | **169.822 (5.566,5181.805)** |
|  | Time to MoCA < 21 | CoxPH | **4.216 (2.426,6.005)** | 0.913 | 4.616 | **<0.001** | **67.730 (11.309,405.640)** |
|  | Time to MoCA < 26 | CoxPH | **2.626 (1.047,4.205)** | 0.806 | 3.260 | **0.001** | **13.818 (2.849,67.011)** |
|  | Time to MDS-UPDRS III Change 5+ | CoxPH | 1.215 (-0.062,2.493) | 0.652 | 1.865 | 0.062 | 3.371 (0.940,12.093) |
| NFL | Time to HY >= 3 | CoxPH | 0.019 (-0.013,0.050) | 0.016 | 1.156 | 0.248 | 1.019 (0.987,1.051) |
|  | Time to Cognitive State Change | CoxPH | 0.020 (-0.027,0.068) | 0.024 | 0.841 | 0.400 | 1.021 (0.973,1.070) |
|  | Time to MoCA < 21 | CoxPH | 0.030 (-0.012,0.071) | 0.021 | 1.397 | 0.163 | 1.030 (0.988,1.074) |
|  | Time to MoCA < 26 | CoxPH | 0.009 (-0.023,0.040) | 0.016 | 0.529 | 0.597 | 1.009 (0.977,1.041) |
|  | Time to MDS-UPDRS III Change 5+ | CoxPH | 0.012 (-0.009,0.034) | 0.011 | 1.137 | 0.256 | 1.013 (0.991,1.035) |
| GFAP | Time to HY >= 3 | CoxPH | 0.003 (-0.001,0.007) | 0.002 | 1.585 | 0.113 | 1.003 (0.999,1.007) |
|  | Time to Cognitive State Change | CoxPH | **0.007 (0.002,0.011)** | 0.002 | 3.069 | **0.002** | **1.007 (1.002,1.011)** |
|  | Time to MoCA < 21 | CoxPH | **0.005 (0.000,0.009)** | 0.002 | 2.082 | **0.037** | **1.005 (1.000,1.009)** |
|  | Time to MoCA < 26 | CoxPH | 0.001 (-0.002,0.005) | 0.002 | 0.680 | 0.496 | 1.001 (0.998,1.005) |
|  | Time to MDS-UPDRS III Change 5+ | CoxPH | 0.002 (-0.001,0.004) | 0.001 | 1.425 | 0.154 | 1.002 (0.999,1.004) |
| Aβ40 plasma | Time to HY >= 3 | CoxPH | -0.001 (-0.006,0.005) | 0.003 | -0.292 | 0.771 | 0.999 (0.994,1.005) |
|  | Time to Cognitive State Change | CoxPH | -0.003 (-0.016,0.010) | 0.007 | -0.394 | 0.693 | 0.997 (0.985,1.010) |
|  | Time to MoCA < 21 | CoxPH | 0.003 (-0.006,0.012) | 0.005 | 0.694 | 0.488 | 1.003 (0.994,1.012) |
|  | Time to MoCA < 26 | AFT | -0.004 (-0.010,0.002) | 0.003 | -1.244 | 0.214 | 1.004 (0.998,1.009) |
|  | Time to MDS-UPDRS III Change 5+ | CoxPH | -0.001 (-0.005,0.003) | 0.002 | -0.565 | 0.572 | 0.999 (0.995,1.003) |
| Aβ42 plasma | Time to HY >= 3 | CoxPH | 0.042 (-0.006,0.090) | 0.025 | 1.710 | 0.087 | 1.043 (0.994,1.094) |
|  | Time to Cognitive State Change | CoxPH | -0.000 (-0.096,0.095) | 0.049 | -0.006 | 0.995 | 1.000 (0.909,1.100) |
|  | Time to MoCA < 21 | CoxPH | 0.038 (-0.028,0.104) | 0.034 | 1.116 | 0.264 | 1.038 (0.972,1.109) |
|  | Time to MoCA < 26 | AFT | -0.019 (-0.067,0.030) | 0.025 | -0.748 | 0.454 | 1.017 (0.972,1.065) |
|  | Time to MDS-UPDRS III Change 5+ | CoxPH | -0.012 (-0.042,0.017) | 0.015 | -0.803 | 0.422 | 0.988 (0.959,1.018) |
| Aβ42/Aβ40 plasma | Time to HY >= 3 | CoxPH | **24.727 (8.799,40.655)** | 8.126 | 3.043 | **0.002** | **5.48E10 (6627.630,4.53E17)** |
|  | Time to Cognitive State Change | CoxPH | 6.569 (-51.090,64.228) | 29.418 | 0.223 | 0.823 | 712.790 (0.000,7.833E27) |
|  | Time to MoCA < 21 | CoxPH | 6.970 (-18.132,32.071) | 12.807 | 0.544 | 0.586 | 1063.776 (0.000,8.481E13) |
|  | Time to MoCA < 26 | AFT | 8.475 (-14.045,30.995) | 11.490 | 0.738 | 0.461 | 0.0004 (0.000,473580.2) |
|  | Time to MDS-UPDRS III Change 5+ | CoxPH | -2.308 (-14.082,9.466) | 6.007 | -0.384 | 0.701 | 0.099 (0.000,12919.11) |

***Cox proportional hazards (CoxPH) or Weibull accelerated failure time (AFT) models were selected automatically by the proportional hazards assumption test. All models adjusted for age and sex. Bold p-values indicate p < 0.05.***

#### Supplement Table 4c: All Continuous HC Progression and Clinical Milestones

| **Biomarker** | **Milestone** | **Model** | **Coefficient (95% CI)** | **SE** | **Z** | ***p*** | **HR (95% CI)** |
| --- | --- | --- | --- | --- | --- | --- | --- |
| **pTau217 plasma** | **Progression Milestones** | | | | | | |
|  | Any progression milestone | CoxPH | -1.4354 (-7.9169,5.0461) | 3.3069 | -0.4341 | 0.6642 | 0.2380 (0.0004,155.4223) |
|  | Walking and balance domain | CoxPH | -3.4213 (-18.2543,11.4118) | 7.5679 | -0.4521 | 0.6512 | 0.0327 (0.0000,90380.88) |
|  | Activities of daily living domain | AFT | 10.5730 (-37.4905,58.6365) | 24.5222 | 0.4312 | 0.6664 | 0.0002 (0.0000,2.078E12) |
|  | **Clinical Milestones** | | | | | | |
|  | Time to MoCA < 21 | CoxPH | -174.082 (-2.307E7,23070758) | 11770884 | -0.0000 | 1.0000 | 0.0000 (NA) |
|  | Time to MoCA < 26 | CoxPH | 3.6378 (1.1483,6.1272) | 1.2701 | 2.8641 | **0.0042** | 38.0074 (3.1529,458.1692) |
|  | Time to MDS-UPDRS III Change 5+ | CoxPH | 2.8938 (-0.2653,6.0529) | 1.6118 | 1.7954 | 0.0726 | 18.0619 (0.7670,425.3591) |
| **NFL** | **Progression Milestones** | | | | | | |
|  | Any progression milestone | AFT | -0.0101 (-0.0616,0.0415) | 0.0263 | -0.3834 | 0.7014 | 1.0164 (0.9349,1.1050) |
|  | Walking and balance domain | CoxPH | -0.0515 (-0.3426,0.2396) | 0.1485 | -0.3469 | 0.7287 | 0.9498 (0.7099,1.2707) |
|  | Activities of daily living domain | AFT | -0.1327 (-0.3597,0.0943) | 0.1158 | -1.1456 | 0.2519 | 1.1289 (0.9446,1.3492) |
|  | **Clinical Milestones** | | | | | | |
|  | Time to MoCA < 21 | CoxPH | -0.3139 (-6007.14,6006.515) | 3064.709 | -0.0001 | 0.9999 | 0.7306 (NA) |
|  | Time to MoCA < 26 | CoxPH | -0.0015 (-0.0447,0.0418) | 0.0221 | -0.0658 | 0.9475 | 0.9985 (0.9563,1.0427) |
|  | Time to MDS-UPDRS III Change 5+ | CoxPH | 0.0064 (-0.0472,0.0601) | 0.0274 | 0.2355 | 0.8139 | 1.0065 (0.9539,1.0619) |
| **GFAP** | **Progression Milestones** | | | | | | |
|  | Any progression milestone | CoxPH | 0.0025 (-0.0080,0.0130) | 0.0053 | 0.4675 | 0.6401 | 1.0025 (0.9920,1.0131) |
|  | Walking and balance domain | CoxPH | 0.0074 (-0.0105,0.0253) | 0.0091 | 0.8070 | 0.4196 | 1.0074 (0.9895,1.0256) |
|  | Activities of daily living domain | AFT | -0.0005 (-0.0333,0.0322) | 0.0167 | -0.0305 | 0.9756 | 1.0004 (0.9740,1.0275) |
|  | **Clinical Milestones** | | | | | | |
|  | Time to MoCA < 21 | CoxPH | -0.0601 (-4208.97,4208.848) | 2147.402 | -0.0000 | 1.0000 | 0.9417 (NA) |
|  | Time to MoCA < 26 | CoxPH | 0.0020 (-0.0045,0.0084) | 0.0033 | 0.5900 | 0.5552 | 1.0020 (0.9955,1.0085) |
|  | Time to MDS-UPDRS III Change 5+ | CoxPH | 0.0027 (-0.0051,0.0105) | 0.0040 | 0.6736 | 0.5006 | 1.0027 (0.9949,1.0106) |

#### Supplement Table 4d: All Categorical sPD Progression Milestones

| **Biomarker** | **Milestone** | **Parameter / Comparison** | **Model** | **Beta (95% CI)** | **SE** | **Z** | **p** | **HR (95% CI)** |
| --- | --- | --- | --- | --- | --- | --- | --- | --- |
| **pTau217 plasma** | Any progression milestone | Overall | AFT |  |  | 2.535 | **0.040** |  |
|  |  | T1 vs T2 | AFT | -0.010 (-0.201,0.181) | 0.097 | -0.102 | 0.919 | 1.016 (0.744,1.388) |
|  |  | T1 vs T3 | AFT | 0.219 (0.021,0.418) | 0.101 | 2.170 | 0.030 | 0.699 (0.504,0.968) |
|  |  | T2 vs T3 | AFT | 0.229 (0.031,0.427) | 0.101 | 2.270 | 0.023 | 0.687 (0.496,0.952) |
|  | Walking and balance domain | Overall | AFT |  |  | 2.084 | 0.114 |  |
|  | Motor complications domain | Overall | CoxPH |  |  | 1.322 | 0.418 |  |
|  | Cognitive domain | Overall | CoxPH |  |  | 3.290 | **0.004** |  |
|  |  | T1 vs T2 | CoxPH | 0.001 (-0.455,0.458) | 0.233 | 0.006 | 0.995 | 1.001 (0.635,1.580) |
|  |  | T1 vs T3 | CoxPH | -0.606 (-1.048,-0.164) | 0.226 | -2.686 | 0.007 | 0.546 (0.351,0.849) |
|  |  | T2 vs T3 | CoxPH | -0.607 (-1.021,-0.193) | 0.211 | -2.875 | 0.004 | 0.545 (0.360,0.824) |
|  | Autonomic dysfunction domain | Overall | CoxPH |  |  | 2.580 | **0.036** |  |
|  |  | T1 vs T2 | CoxPH | 0.223 (-0.261,0.707) | 0.247 | 0.903 | 0.366 | 1.250 (0.770,2.028) |
|  |  | T1 vs T3 | CoxPH | -0.371 (-0.836,0.093) | 0.237 | -1.567 | 0.117 | 0.690 (0.433,1.098) |
|  |  | T2 vs T3 | CoxPH | -0.594 (-1.054,-0.135) | 0.234 | -2.536 | 0.011 | 0.552 (0.349,0.874) |
|  | Functional dependence domain | Overall | AFT |  |  | 2.480 | **0.046** |  |
|  |  | T1 vs T2 | AFT | -0.102 (-0.305,0.101) | 0.104 | -0.983 | 0.326 | 1.219 (0.822,1.807) |
|  |  | T1 vs T3 | AFT | 0.153 (-0.046,0.352) | 0.102 | 1.508 | 0.132 | 0.743 (0.504,1.095) |
|  |  | T2 vs T3 | AFT | 0.255 (0.051,0.459) | 0.104 | 2.455 | 0.014 | 0.609 (0.410,0.906) |
| **NFL** | Any progression milestone | Overall | CoxPH |  |  | 2.213 | 0.086 |  |
|  | Walking and balance domain | Overall | AFT |  |  | 0.872 | 0.684 |  |
|  | Motor complications domain | Overall | CoxPH |  |  | 0.766 | 0.745 |  |
|  | Cognitive domain | Overall | CoxPH |  |  | 2.642 | **0.031** |  |
|  |  | T1 vs T2 | CoxPH | -0.160 (-0.651,0.331) | 0.251 | -0.640 | 0.522 | 0.852 (0.521,1.392) |
|  |  | T1 vs T3 | CoxPH | -0.647 (-1.189,-0.105) | 0.277 | -2.340 | 0.019 | 0.524 (0.304,0.900) |
|  |  | T2 vs T3 | CoxPH | -0.487 (-0.912,-0.062) | 0.217 | -2.244 | 0.025 | 0.615 (0.402,0.940) |
|  | Autonomic dysfunction domain | Overall | CoxPH |  |  | 1.211 | 0.480 |  |
|  | Functional dependence domain | Overall | AFT |  |  | 1.569 | 0.292 |  |
| **GFAP** | Any progression milestone | Overall | CoxPH |  |  | 0.283 | 0.961 |  |
|  | Walking and balance domain | Overall | AFT |  |  | 1.401 | 0.375 |  |
|  | Motor complications domain | Overall | CoxPH |  |  | 2.060 | 0.120 |  |
|  | Cognitive domain | Overall | CoxPH |  |  | 2.236 | 0.082 |  |
|  | Autonomic dysfunction domain | Overall | CoxPH |  |  | 1.322 | 0.417 |  |
|  | Functional dependence domain | Overall | AFT |  |  | 1.158 | 0.512 |  |
| **Aβ40 plasma** | Any progression milestone | Overall | CoxPH |  |  | 0.580 | 0.845 |  |
|  | Walking and balance domain | Overall | AFT |  |  | 0.744 | 0.758 |  |
|  | Motor complications domain | Overall | CoxPH |  |  | 1.747 | 0.217 |  |
|  | Cognitive domain | Overall | CoxPH |  |  | 0.843 | 0.701 |  |
|  | Autonomic dysfunction domain | Overall | CoxPH |  |  | 0.498 | 0.883 |  |
|  | Functional dependence domain | Overall | CoxPH |  |  | 0.536 | 0.866 |  |
| **Aβ42 plasma** | Any progression milestone | Overall | CoxPH |  |  | 1.116 | 0.536 |  |
|  | Walking and balance domain | Overall | AFT |  |  | 1.564 | 0.294 |  |
|  | Motor complications domain | Overall | CoxPH |  |  | 0.343 | 0.943 |  |
|  | Cognitive domain | Overall | CoxPH |  |  | 1.198 | 0.488 |  |
|  | Autonomic dysfunction domain | Overall | CoxPH |  |  | 1.437 | 0.356 |  |
|  | Functional dependence domain | Overall | CoxPH |  |  | 0.380 | 0.930 |  |
| **Aβ42/ Aβ40 plasma** | Any progression milestone | Overall | CoxPH |  |  | 1.317 | 0.420 |  |
|  | Walking and balance domain | Overall | AFT |  |  | 1.532 | 0.309 |  |
|  | Motor complications domain | Overall | CoxPH |  |  | 0.418 | 0.916 |  |
|  | Cognitive domain | Overall | CoxPH |  |  | 1.705 | 0.234 |  |
|  | Autonomic dysfunction domain | Overall | CoxPH |  |  | 1.461 | 0.344 |  |
|  | Functional dependence domain | Overall | CoxPH |  |  | 1.019 | 0.595 |  |

#### Supplement Table 4e: All Categorical sPD Clinical Milestones

| **Biomarker** | **Clinical Milestone** | **Model** | **Parameter / Comparison** | **Beta (95% CI)** | **SE** | **Z** | **p** | **HR (95% CI)** |
| --- | --- | --- | --- | --- | --- | --- | --- | --- |
| **pTau217 plasma** | Time to HY >= 3 | AFT | Overall |  |  | 1.476 | 0.337 |  |
|  | Time to Cognitive State Change | AFT | Overall |  |  | 1.074 | 0.562 |  |
|  | Time to MoCA < 21 | CoxPH | Overall |  |  | 3.368 | **0.003** |  |
|  |  | CoxPH | T1 vs T2 | 0.070 (-0.720,0.860) | 0.403 | 0.173 | 0.862 | 1.072 (0.487,2.363) |
|  |  | CoxPH | T1 vs T3 | -0.877 (-1.573,-0.181) | 0.355 | -2.470 | **0.014** | 0.416 (0.207,0.834) |
|  |  | CoxPH | T2 vs T3 | -0.947 (-1.582,-0.312) | 0.324 | -2.922 | **0.003** | 0.388 (0.206,0.732) |
|  | Time to MoCA < 26 | CoxPH | Overall |  |  | 2.205 | 0.088 |  |
|  | Time to MDS-UPDRS III Change 5+ | AFT | Overall |  |  | 2.877 | **0.016** |  |
|  |  | AFT | T1 vs T2 | 0.332 (0.096,0.568) | 0.120 | 2.754 | **0.006** | 0.667 (0.498,0.892) |
|  |  | AFT | T1 vs T3 | 0.270 (0.019,0.521) | 0.128 | 2.112 | **0.035** | 0.719 (0.528,0.979) |
|  |  | AFT | T2 vs T3 | -0.062 (-0.304,0.180) | 0.123 | -0.499 | 0.618 | 1.078 (0.802,1.449) |
| **NFL** | Time to HY >= 3 | CoxPH | Overall |  |  | 1.462 | 0.344 |  |
|  | Time to Cognitive State Change | CoxPH | Overall |  |  | 1.220 | 0.475 |  |
|  | Time to MoCA < 21 | CoxPH | Overall |  |  | 2.680 | **0.028** |  |
|  |  | CoxPH | T1 vs T2 | -0.725 (-1.640,0.191) | 0.467 | -1.552 | 0.121 | 0.485 (0.194,1.210) |
|  |  | CoxPH | T1 vs T3 | -1.272 (-2.244,-0.301) | 0.496 | -2.567 | **0.010** | 0.280 (0.106,0.740) |
|  |  | CoxPH | T2 vs T3 | -0.548 (-1.163,0.067) | 0.314 | -1.745 | 0.081 | 0.578 (0.313,1.070) |
|  | Time to MoCA < 26 | CoxPH | Overall |  |  | 1.047 | 0.578 |  |
|  | Time to MDS-UPDRS III Change 5+ | AFT | Overall |  |  | 0.669 | 0.799 |  |
| **GFAP** | Time to HY >= 3 | CoxPH | Overall |  |  | 0.982 | 0.617 |  |
|  | Time to Cognitive State Change | CoxPH | Overall |  |  | 2.062 | 0.119 |  |
|  | Time to MoCA < 21 | CoxPH | Overall |  |  | 1.352 | 0.401 |  |
|  | Time to MoCA < 26 | CoxPH | Overall |  |  | 1.626 | 0.267 |  |
|  | Time to MDS-UPDRS III Change 5+ | CoxPH | Overall |  |  | 1.784 | 0.204 |  |
| **Aβ40 plasma** | Time to HY >= 3 | CoxPH | Overall |  |  | 0.807 | 0.722 |  |
|  | Time to Cognitive State Change | CoxPH | Overall |  |  | 1.397 | 0.377 |  |
|  | Time to MoCA < 21 | CoxPH | Overall |  |  | 1.380 | 0.386 |  |
|  | Time to MoCA < 26 | AFT | Overall |  |  | 1.228 | 0.470 |  |
|  | Time to MDS-UPDRS III Change 5+ | CoxPH | Overall |  |  | 0.899 | 0.667 |  |
| **Aβ42 plasma** | Time to HY >= 3 | AFT | Overall |  |  | 1.437 | 0.356 |  |
|  | Time to Cognitive State Change | CoxPH | Overall |  |  | 1.556 | 0.298 |  |
|  | Time to MoCA < 21 | CoxPH | Overall |  |  | 0.792 | 0.731 |  |
|  | Time to MoCA < 26 | AFT | Overall |  |  | 1.435 | 0.357 |  |
|  | Time to MDS-UPDRS III Change 5+ | CoxPH | Overall |  |  | 1.284 | 0.439 |  |
| **Aβ42/Aβ40 plasma** | Time to HY >= 3 | CoxPH | Overall |  |  | 1.764 | 0.211 |  |
|  | Time to Cognitive State Change | CoxPH | Overall |  |  | 1.100 | 0.546 |  |
|  | Time to MoCA < 21 | CoxPH | Overall |  |  | 2.067 | 0.118 |  |
|  | Time to MoCA < 26 | AFT | Overall |  |  | 1.759 | 0.213 |  |
|  | Time to MDS-UPDRS III Change 5+ | CoxPH | Overall |  |  | 0.995 | 0.610 |  |

### Supplement Table 4f: All Categorical HC NSD- Progression and Clinical Milestones

| **Biomarker** | **Analysis Type** | **Milestone** | **Model** | **Parameter / Comparison** | **N** | **Events** | **Censored** | **Z** | **p** |
| --- | --- | --- | --- | --- | --- | --- | --- | --- | --- |
| pTau217 plasma | Progression | Any progression milestone | AFT | Overall | 94 | 8 | 86 | 0.628 | 0.821 |
| pTau217 plasma | Clinical | Time to MoCA < 26 | CoxPH | Overall | 97 | 41 | 56 | 1.462 | 0.343 |
| pTau217 plasma | Clinical | Time to MDS-UPDRS III Change 5+ | CoxPH | Overall | 97 | 23 | 74 | 1.756 | 0.214 |
| NFL | Progression | Any progression milestone | CoxPH | Overall | 94 | 8 | 86 | 0.321 | 0.950 |
| NFL | Progression | Walking and balance domain | CoxPH | Overall | 95 | 3 | 92 | 1.604 | 0.276 |
| NFL | Clinical | Time to MoCA < 26 | CoxPH | Overall | 97 | 41 | 56 | 0.466 | 0.897 |
| NFL | Clinical | Time to MDS-UPDRS III Change 5+ | CoxPH | Overall | 97 | 23 | 74 | 0.966 | 0.627 |
| GFAP | Progression | Any progression milestone | CoxPH | Overall | 94 | 8 | 86 | 1.614 | 0.272 |
| GFAP | Clinical | Time to MoCA < 26 | CoxPH | Overall | 97 | 41 | 56 | 0.751 | 0.754 |
| GFAP | Clinical | Time to MDS-UPDRS III Change 5+ | CoxPH | Overall | 97 | 23 | 74 | 1.207 | 0.482 |
